# Butyrate-producing gut bacteria are associated with protection from allergic symptoms after Hurricane Harvey

**DOI:** 10.1101/2021.12.09.21267553

**Authors:** Kristen M. Panthagani, Kristi L. Hoffman, Abiodun Oluyomi, Jesus Sotelo, Christopher Stewart, Georgina Armstrong, Dan Na Luo, Melissa Bondy, Cheryl Lyn Walker, Joseph F. Petrosino

**Author notes:** **Materials and Correspondence** Correspondence should be addressed to Dr. Joseph Petrosino and Dr. Cheryl Walker.

## Abstract

Hurricane Harvey caused record-breaking, catastrophic flooding across the city of Houston. After floodwaters receded, several health concerns arose, including the potential adverse impact of exposure to mold in flooded homes. We rapidly launched the Houston Hurricane Harvey Health Study to evaluate if microbiome sampling in the wake of a disaster could inform flood-associated environmental exposures and adverse health outcomes. We enrolled a total of 347 subjects at 1-month and 12-months post-Harvey, collecting human (stool, nasal, saliva) and environmental (house swab) samples to profile the bacterial and fungal microbiota. Here we show reported exposure to mold was associated with increased risk of allergic symptoms for up to one year post-disaster, and that butyrate-producing bacteria in the gut were linked to protection from allergic symptoms in mold-exposed individuals. Together, these data provide new insights into how microbiome:environment interactions may influence health in the setting of a flood-related disaster.

## Introduction

On August 25, 2017, Hurricane Harvey made landfall in Rockport, Texas as a category 4 hurricane, marking the beginning of the second costliest tropical cyclone disaster in US history.^1^ Over the next several days, the slow-moving storm system stalled over Harris County, dropping over 50 inches of rain, breaking the record for continental US rainfall.^2^ Catastrophic flooding across the greater Houston area impacted over 200,000 homes in Harris County alone.^3^ Clean-up efforts lasted months, with many individuals “mucking and gutting” their own homes. Given the unprecedented levels of flooding, extensive clean-up efforts, and the humid climate in Southeast Texas, exposure to mold and other environmental microbes was a significant health concern.^4^

Mold can cause a variety of health problems, including sequelae of mycotoxin ingestion and invasive infections in immunocompromised individuals.^5^ However, for healthy individuals in non-occupational settings, the primary concern is allergic response and/or respiratory tract irritation caused by inhaled mold spores.^5,6^ Mold growth and/or indoor dampness are associated with a variety of allergic pathologies including allergic rhinitis, eczema, and asthma exacerbation.^7^ As many Houstonians remained in flood-damaged homes, the health effects of mold exposure became a significant concern.^8^ Previous studies on the health impact of mold growth caused by hurricanes have been inconclusive: after Hurricanes Katrina and Rita, 46% of homes assessed in New Orleans had documented mold growth, but no increase in symptoms related to mold exposure was detected.^6^ However, these studies were limited to individuals who sought medical care and likely overlooked those who chose not to seek treatment or lacked access to healthcare.^6^ Thus, the health impact of mold exposure after flooding events remains unknown, particularly in communities with limited resources.

The role of the microbiome in regulating allergic immune responses is becoming increasingly apparent.^9^ Microbial-derived metabolites such as butyrate, a short-chain fatty acid (SCFA) derived from bacterial fermentation of dietary fiber, have been shown to directly regulate eosinophil migration and survival and reduce allergic airway inflammation.^10,11^ Given these findings, we hypothesized that human microbiome composition may be associated with health outcomes following exposure to environmental allergens such as mold.

We launched the Houston Hurricane Harvey Health (Houston3H) Study to assess environmental exposures and health outcomes of Houstonians impacted by Hurricane Harvey. We found mold exposure was significantly associated with increased risk of at least one allergic symptom up to a year after Hurricane Harvey. Furthermore, we discovered that subjects with higher levels of butyrate-producing gut bacteria were at lower risk for allergic symptoms even with mold exposure. Therefore, while exposure to mold in the setting of a flood-related disaster can increase risk of allergic symptoms, the gut microbiome may play an important role in mitigating this risk.

## Results

### Cohort Characteristics

The Houston3H Study enrolled a total of 347 subjects from 270 households across two timepoints: 206 subjects (178 households) at 1-month post-Harvey and 266 subjects (199 households) at 12-months post-Harvey, which includes 125 subjects (107 households) who participated at both time points. Subjects were recruited from four distinct Houston-area neighborhoods impacted by Hurricane Harvey: Addicks, Baytown, East Houston, and Bellaire/Meyerland. Demographics varied significantly by neighborhood: the majority of subjects from Addicks and Bellaire/Meyerland were Non-Hispanic White and lived in census tracts with lower Area Deprivation Index (ADI) scores, indicative of higher socioeconomic status, while the majority of subjects from Baytown and East Houston were Hispanic or Non-Hispanic Black and had higher ADI scores (**Figure 1a, Supplementary Table 1**). Details of this cohort have been previously described.^12^

**Figure 1.**
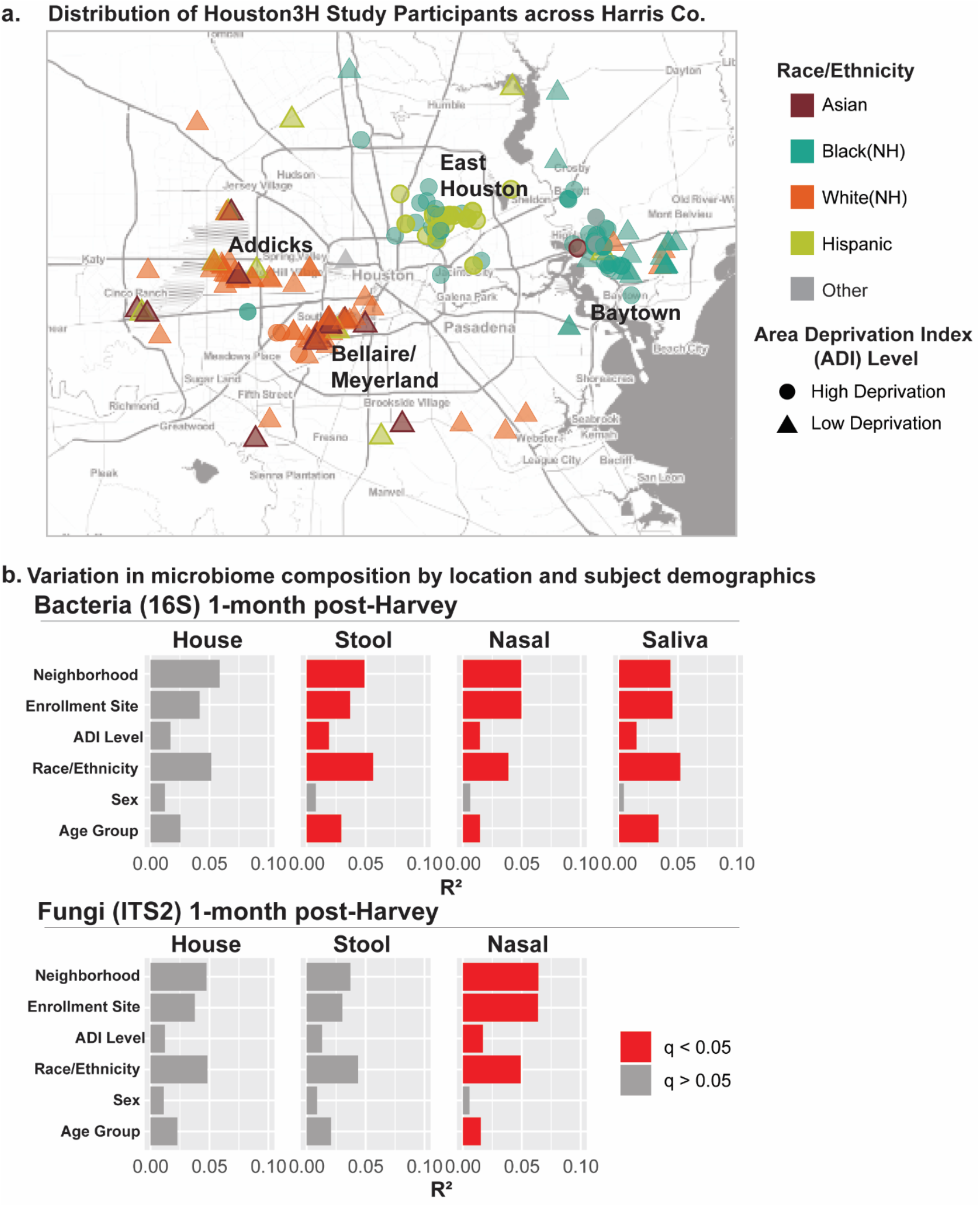
Impact of subject demographics on bacterial and fungal microbiome composition. **a**. Distribution of study participants (1-month post-Harvey) across Houston-area neighborhoods. Color indicates race/ethnicity of subjects, shape indicates census tract-level calculation of area deprivation index (ADI) of subjects’ homes, with higher ADI scores indicative of lower socioeconomic status. Placement of markers on map represent approximate location of participants’ homes at the time of Hurricane Harvey. **b**. Principal coordinate analysis (PCoA) of Binary Jaccard distances was used to evaluate associations between demographic variables and microbiome composition at 1-month post-Harvey. PERMANOVA was used to calculate R^2^ (percent of total microbial variation accounted for by each variable) and p-values. Bonferroni correction of p-values was used to calculate q-values for each body site. Total samples (n) remaining in each analysis after rarefaction: house 16S (n=80), nasal 16S (n=186), stool 16S (n=120), saliva 16S (n=194), house ITS2 (n=93), nasal ITS2 (n=189), stool ITS2 (n=102). Only one house swab per household was included in house microbiome analyses. Sample numbers and statistics for each individual analysis are provided in Supplementary Table 4.

### Human and house microbiome composition varies by demographics and geography

We first evaluated the microbiome composition in association with subject demographics and geography. We profiled the bacterial microbiome using 16S-V4 rRNA amplicon sequencing, and beta diversity (Binary Jaccard) analyses revealed strong associations between race/ethnicity, age, neighborhood, enrollment site, and ADI and the human microbiome (**Figure 1b, Supplementary Figure 1)**. The stool and salivary microbiomes were more strongly associated with race/ethnicity than any other demographic variable tested, suggesting that race and ethnicity and/or associated collinear cultural and dietary factors are major drivers of microbial ecology in these body sites. In contrast, the nasal microbiome was most strongly associated with neighborhood, suggesting that geographic location plays a significant role in shaping the nasal microbiome.

Using Internal Transcribed Spacer-2 (ITS2) amplicon sequencing, we profiled the fungal microbiome (mycobiome) of nasal, stool and house swab samples. Beta diversity analyses revealed the same variables that drove nasal bacterial microbiome composition impacted the nasal mycobiome, with neighborhood more strongly associated with mycobiome composition than race/ethnicity. However, unlike the stool bacterial microbiome, we found no association between the taxonomic composition of the stool mycobiome and any demographic variables tested. We additionally profiled a snapshot of the microbes in the home environment (house microbiome). Subjects sampled their homes by swabbing the entry/threshold of their home’s front door. At 1-month post-Harvey (**Figure 1b**), beta diversity analyses revealed no association between the house microbiome (fungal and bacterial) and demographic variables. At 12-months post-Harvey, we found a significant association between geographical variables and the house microbiome (both fungal and bacterial), suggesting that geographical location (neighborhood) plays a significant role in shaping the community of microbes in the home environment (**Supplementary Figure 1**).

### Impact of mold exposure on allergic symptoms

We next assessed the relationship between allergic symptoms and mold exposure. Our questionnaire was designed using the NIH Disaster Research Response Resources (https://dr2.nlm.nih.gov) and asked subjects to report symptoms known to be associated with indoor dampness or mold exposure^7,13^ including throat irritation, sinus irritation, eye irritation, wheezing, cough, shortness of breath, and skin rash (both time points) and worsening asthma (1-month post-Harvey only). For simplicity, we collectively refer to this group of mold-associated symptoms as “allergic symptoms.” For the seven individual symptoms included at both time points, our questionnaire asked subjects to report symptoms that occurred following the hurricane, excluding those resulting from a cold or seasonal allergies and inclusive of symptoms that had since resolved. The majority of participants reported at least one allergic symptom (70.4% in 1-month post-Harvey cohort, 65.0% in 12-months post-Harvey cohort), with minimal variation in subject demographics by allergic symptoms (**Supplementary Tables 2-3**). Building on our prior work assessing the relationship between hurricane exposures and individual allergic symptoms,^12^ in the present study we analyzed symptoms collectively and assessed the relationship between reported mold exposure and the presence of at least one allergic symptom. At 1-month post-Harvey, both reported exposure to visible mold and new signs of mold in the home were associated with increased risk of at least one allergic symptom (**Figure 2**). In the 12-months post-Harvey cohort, when the majority of flood remediation was complete, only signs of mold growth in the home was associated with increased risk of allergic symptoms (**Figure 2**).

**Figure 2.**
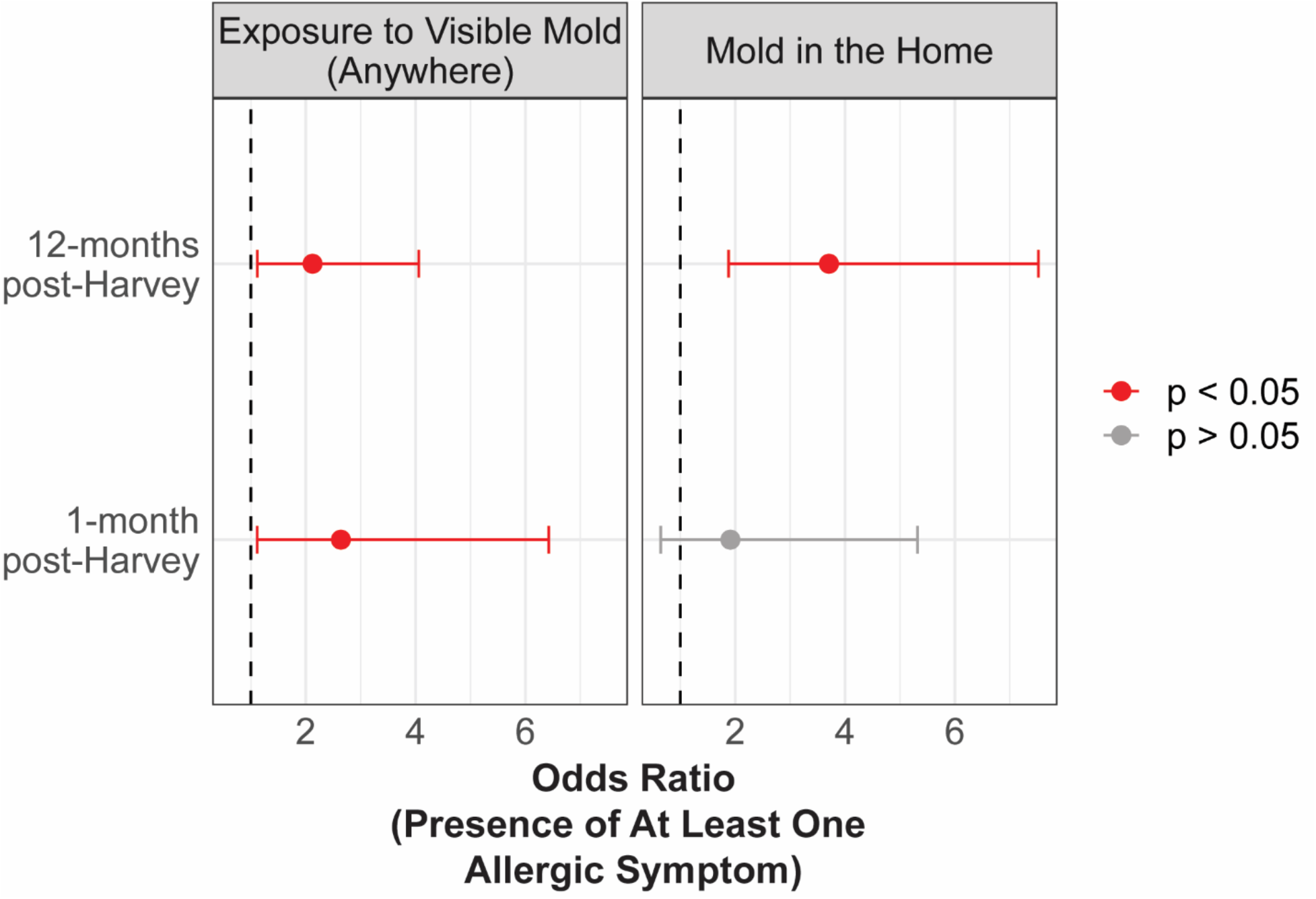
Association between reported exposure to mold and presence of allergic symptoms. Odds ratios calculated using reported exposures to mold (visible mold anywhere, or mold in the home) and presence of at least one allergic symptom at each time point, controlling for race/ethnicity, age, sex, and education level. Circles indicate odds ratios and bars indicate 95% confidence intervals. Total subjects in each analysis, excluding missing values: exposure to visible mold, 1-month post-Harvey (n = 168), exposure to visible mold, 12-months post-Harvey (n= 243), mold in the home, 1-month post-Harvey (n = 151), mold in the home, 12-months post-Harvey (n = 207).

### Gut bacterial microbiome is associated with allergic symptoms when mold is present in the home

As the microbiome has a major influence on systemic immune responses to allergens,^14^ we asked if there was an association between reported allergic symptoms and the microbiome. Using 16S-V4 and ITS2 amplicon sequencing, we discovered a strong association between the gut bacterial microbiome and allergic symptoms at 1-month post-Harvey, but no association in any other sample type (**Figure 3a**). We performed Whole Genome Shotgun (WGS) sequencing to verify this association using species-level resolution of the gut microbiome, and again found significant clustering of taxonomic composition by presence of allergic symptoms (**Figure 3b**). This association did not persist at 12-months post-Harvey in the gut or any other sample type (**Supplementary Figure 2**). Among individuals who reported new signs of mold in their home after Harvey, we found that gut microbiome composition at 12-months post-Harvey was associated with allergic symptoms, potentially due to the ongoing allergen exposure even after remediation efforts (**Figure 3b**). No significant differences in gut microbiome alpha diversity (species richness) by allergic symptoms were detected (**Figure 3c)**.

**Figure 3.**
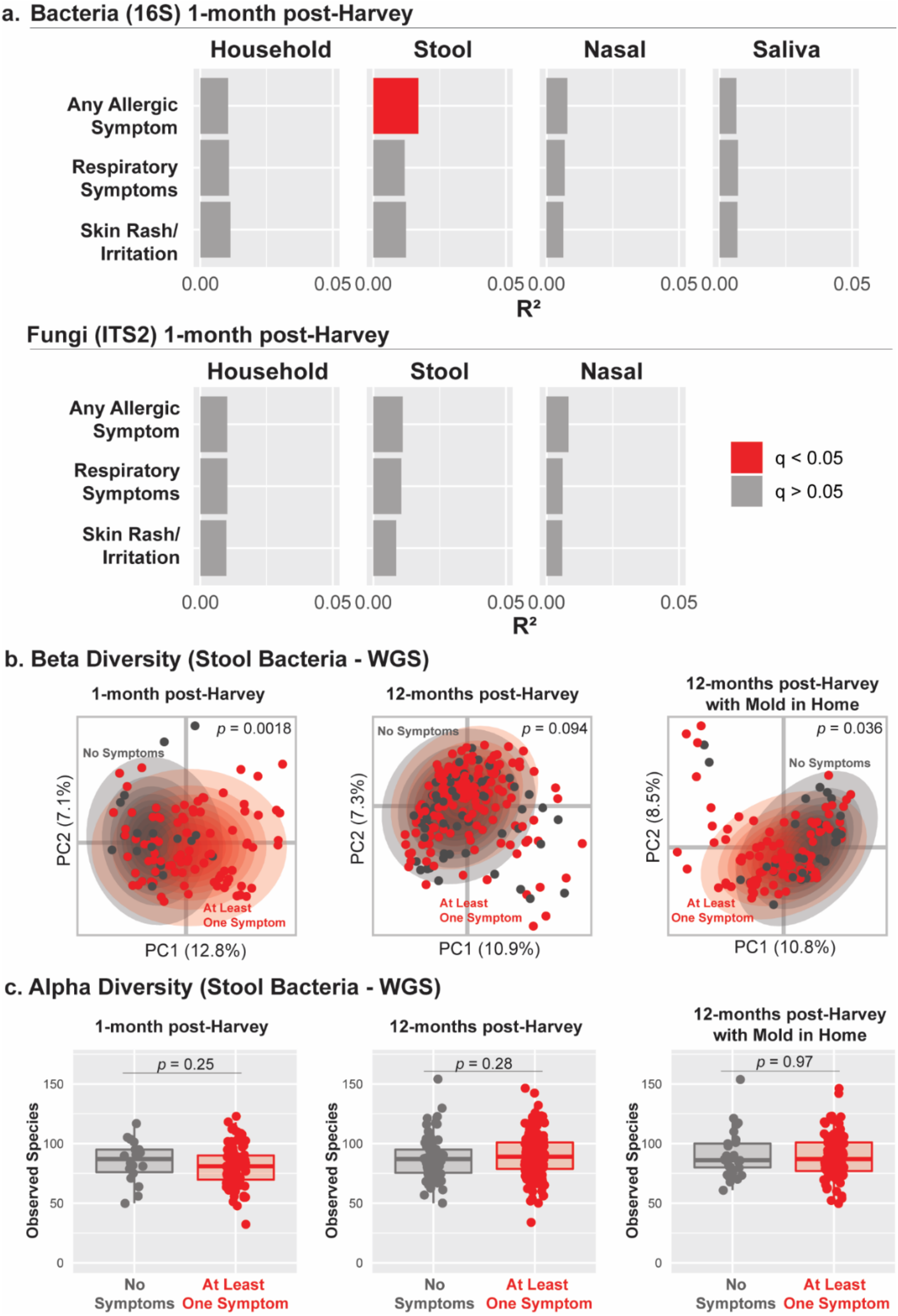
Association between microbiome composition and allergic health outcomes. **a**. PCoA analyses of Binary Jaccard distances were used to evaluate associations between bacterial (16S) and fungal (ITS2) microbiota composition and reported allergic symptoms within 1-month post-Harvey. PERMANOVA was used to calculate R^2^ (percent of total microbial variation accounted for by each variable) and p-values, followed by Bonferroni correction. Total samples in each analysis after rarefaction: house 16S (n=77), stool 16S (n=107), nasal 16S (n=161), saliva 16S (n=165), house ITS2 (n=92), stool ITS2 (n=89), nasal ITS2 (n=162). Sample numbers and statistics for each individual analysis are provided in Supplementary Table 4. **b**. PCoA analysis of bacterial species profiled by Whole Genome Shotgun (WGS) sequencing of stool samples evaluating association between gut bacterial species and allergic symptoms at 1-month post-Harvey (n=18 without symptoms, n=88 with symptoms, PERMANOVA *p* = 0.0018 and R^2^ = 0.021, PERMDISP *p* < 0.001) and 12-months post-Harvey (n=67 without symptoms, n = 156 with symptoms, PERMANOVA *p* = 0.094 and R^2^ = 0.006, PERMDISP *p* = 0.52) in all subjects, and sub-analysis of 12-months post-Harvey subjects with reported signs of mold in the home (n=29 without symptoms, n=97 with symptoms, PERMANOVA *p* = 0.036 and R^2^ = 0.012, PERMDISP *p* = 0.20). Outer ellipse represents 95% normal confidence ellipse for each group. **c**. No difference in gut bacterial alpha diversity was detected at any time point (two-sided Mann-Whitney test; *p* > 0.05). Box plots indicate median and interquartile range (IQR), whiskers show smallest (lower whisker) or largest (upper whisker) value within 1.5 times the IQR.

### Butyrate-producing bacteria are enriched in the gut microbiome of subjects without allergic symptoms

We next determined which bacterial species were driving the observed gut microbiome:environmental mold interaction with allergic health outcomes. Linear Discriminant Analysis Effect Size^15^ (LEfSe) revealed that species enriched in subjects without allergic symptoms had the common ability to produce butyrate, a microbial product known to regulate immune cells and mitigate allergic responses^11,16^ (**Figure 4a,b**). At 1-month post-Harvey, multiple butyrate-producing species were enriched in subjects without allergic symptoms, including *Roseburia intestinalis, Roseburia inulinovorans, Bacteroides finegoldii, Coprococcus comes*, and *Eubacterium ventriosum*. At 12-months post-Harvey, only a single butyrate-producing bacterial species (*Bacteroides faecis)* was enriched in subjects without allergic symptoms. However, co-occurrence analysis revealed that *B. faecis* frequently co-occurred with other butyrate-producing bacteria including multiple *Alistipes, Bacteroides*, and *Roseburia* spp. (**Figure 4c,d**). This suggests that butyrate production may be driven by a cooperative network of multiple bacteria rather than dominance of only a few species driving butyrate production. Notably, *Ruminococcus gnavus*, which negatively co-occurs with many butyrate-producing bacteria, was enriched in subjects with allergic symptoms at 12-months post-Harvey. This pattern of *R. gnavus* negative co-occurrence was remarkably consistent across both time points as well as multiple independent cohorts from the US and Europe (**Supplementary Figure 3**).

**Figure 4.**
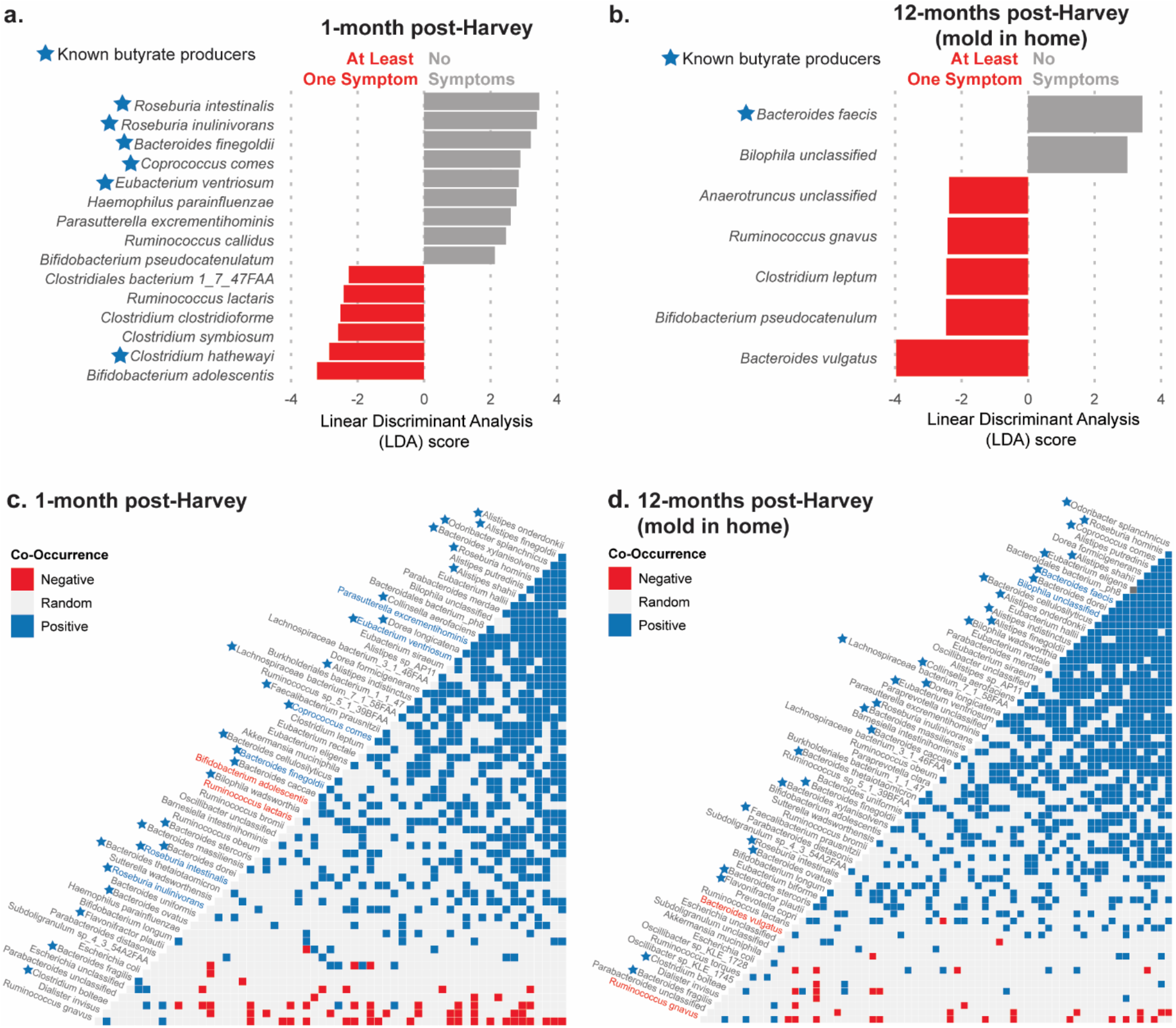
Gut bacterial species co-occurrence and association with allergic health outcomes. Linear discriminant analysis Effect Size (LEfSe) reveals bacterial species associated with absence of allergic symptoms (gray) and presence of allergic symptoms (red) at 1-month post-Harvey (**a**, n=18 without symptoms, n=88 with symptoms) and 12-months post-Harvey for subjects with signs of mold in the home (**b**, n=29 without symptoms, n=97 with symptoms) (Kruskal-Wallis *p* < 0.05, LDA Score > 2.0, species prevalence > 20% of samples). Co-occurrence analysis of all bacterial species reveals positive (blue) and negative (red) associations between gut bacterial species in subjects at 1-month post-Harvey (**c**, n=117) and 12-months post-Harvey with mold in home (**d**, n=130). Species associated with presence and absence of allergic symptoms are highlighted in red and blue, respectively, and blue stars indicate butyrate-producing bacteria. Co-occurrence analyses include species present at >0.1% relative abundance in at least 20% of samples.

### Microbial butyrate metabolism genes are enriched in the gut microbiome of subjects without allergic symptoms

We next examined the functional capacity of the gut microbiome using WGS sequencing data. We found that butyrate metabolism genes were significantly enriched in the guts of subjects without allergic symptoms at 1-month post-Harvey (**Figure 5a**). At 12-months post-Harvey, similar to taxonomic composition (**Figure 3b**), we found that while there was no difference in butyrate metabolic capacity by allergic symptoms across all subjects (**Figure 5b**), when including only subjects reporting signs of mold growth in their home post-Harvey, we saw a significant enrichment of butyrate metabolism genes in the gut microbiome of subjects without allergic symptoms (**Figure 5c**). To test if the consistency of this association was driven primarily by longitudinal subjects who participated at both time points, we next repeated this analysis excluding longitudinal subjects, and found that this association was reproducible in the cohort of subjects who participated at the 12-month post-Harvey time point only (**Supplementary Figure 4**). Given that many bacterial species capable of producing butyrate are also able to produce other short chain fatty acids (SCFA; e.g., propionate and acetate), we asked if gene content of the gut microbiome was enriched for these metabolic pathways, but found no difference between subjects with and without allergic symptoms at either time point (**Figure 5**). To test if additional bacterial functions could also be associated with allergic health outcomes, we performed LEfSe analysis on all mapped functional pathways (**Supplementary Figure 5**), which revealed several other functional pathways associated with allergic health outcomes; notably, fatty acid biosynthesis was enriched in subjects without allergic symptoms at both 1-month and 12-months post-Harvey. Finally, we evaluated factors known to associate with gut microbiome composition including body mass index (BMI)^17^ and recent antibiotic use,^18^ and we found no association between these variables and butyrate metabolism gene abundance at either time point (**Supplementary Figure 6**).

**Figure 5.**
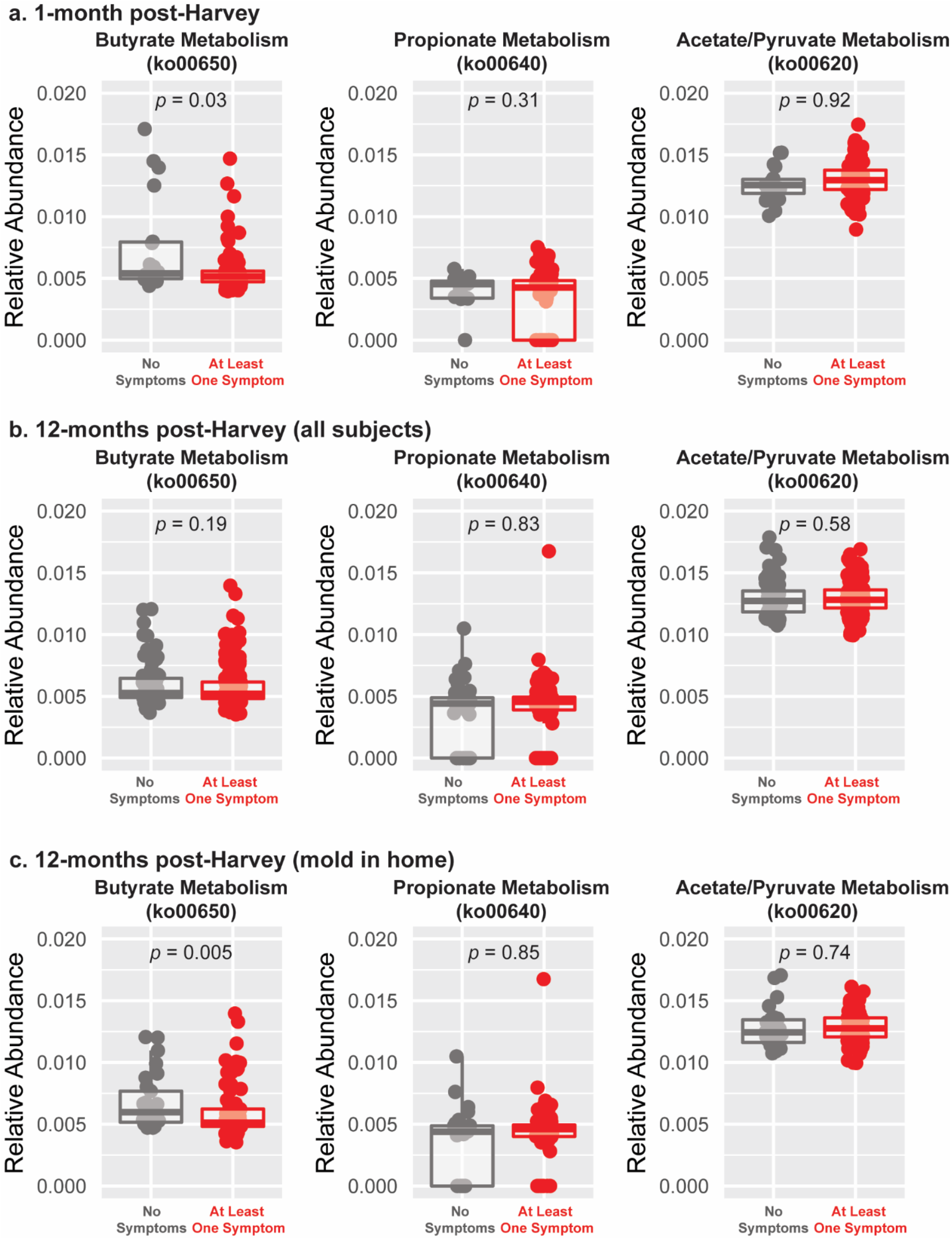
Relative abundance of bacterial genes involved in short-chain fatty acid (SCFA) metabolism in association with allergic symptoms. WGS sequencing was used to determine the relative abundance of metabolic pathways involved in the production of the three major SCFAs (butyrate, propionate, and acetate) at 1-month post-Harvey (**a**, n=18 without symptoms, n=88 with symptoms), 12-months post-Harvey (all subjects) (**b**, n=67 without symptoms, n=156 with symptoms) and 12-months post-Harvey (mold in home only) (**c**, n=29 without symptoms, n=97 with symptoms). All statistical tests reported are one-sided Mann-Whitney tests. Box plots indicate median and interquartile range (IQR), whiskers show smallest (lower whisker) or largest (upper whisker) value within 1.5 times the IQR.

### Impact of hurricane exposures on microbiome composition

We next sought to determine if hurricane exposures may have altered human microbiome composition. Beta diversity analysis revealed no significant associations between hurricane exposures (exposure to dirty water, sewage, visible mold, new signs of mold growth in the home since Harvey, and whether subjects were rescued or involved in clean-up work) and human microbiome composition in any sample type (**Supplementary Figure 7**). We next evaluated if ITS2 amplicon sequencing of house threshold samples can be used to assess mold growth in the home, and discovered no significant association between house threshold mycobiome profiles and subject reported mold growth. (**Supplementary Figure 7**). Given the variation in the mycobiome across neighborhoods, we hypothesized this lack of association may be explained by neighborhood-specific mold signatures, supposing a single signature indicative of mold growth may not be universal across Houston. At 12-months post-Harvey, we found that each subjects’ nasal mycobiome was more similar (shorter Binary Jaccard distance) to the mycobiome of their own home relative to other homes, suggesting each home has its own unique mold exposure profile. However, the similarity (Binary Jaccard distance) between a subjects’ nasal and house mycobiome was not associated with reported exposure to mold or signs of mold growth in the home (**Supplementary Figure 8**).

## Discussion

By rapidly launching the Houston3H study following an unprecedented flooding disaster in our city, we were able to assemble a uniquely diverse cohort to evaluate the interaction between environmental exposures and microbiome composition in a disaster setting. We found significant associations between exposure to mold and allergic symptoms. At 1-month post-Harvey, both reported exposure to visible mold (anywhere) and reported signs of mold growth in the home were associated with increased risk of at least one allergic symptom, while at 12-months post-Harvey only signs of mold in the home was associated with increased risk of allergic symptoms. These data suggest that at 1-month post-Harvey, mold in the home was not necessarily the primary source of mold exposure, while months after the Hurricane, mold growth in the home likely became the primary source.

Our study provides epidemiological corroboration of prior observational and mechanistic work establishing a strong link between butyrate and protection against allergic inflammation. Butyrate, a SCFA derived from fermentation of dietary fiber by gut microbiota,^10^ is a key microbial product linking the microbiome and the immune system via regulation of inflammatory cytokine production and induction of regulatory T cells.^16^ Multiple studies have found higher levels of butyrate-producing bacteria and/or fecal levels of SCFAs to be associated with protection against allergic health outcomes including asthma, allergic rhinitis, atopic dermatitis, and allergic sensitization.^19–22^ Several interventional studies in animal models have established a causal link between dietary fiber intake, butyrate, and protection against allergic airway disease including asthma.^11,20,23,24^ Butyrate directly inhibits eosinophil migration, adhesion, and survival *in vitro*, and studies in adults with stable asthma have shown that consumption of fiber (with or without probiotics) acutely decreases airway inflammation and improves asthma control.^11,25,26^ Together, these observations provide strong evidence that butyrate derived from bacterial fermentation of dietary fiber plays a significant role in attenuating allergic immune responses.

In our study, the association between lower levels of bacterial butyrate metabolism genes and presence of allergic symptoms was found at 1-month post-Harvey in all subjects, while at 12-months post-Harvey, this association was only detected in subjects who reported new signs of mold growth in their home after the hurricane. This pattern suggests a possible microbiome:environment interaction, whereby lack of butyrate producers in the gut may prime individuals for an allergic response, but individuals will only become symptomatic with antigen exposure. A sub-analysis excluding longitudinal subjects verified this association is reproducible in two distinct cohorts of participants.

Given that the gut microbiome represents a cooperative network of microbes rather than individual species exerting functional effects in isolation, it is possible that a gut microbial ecosystem marked by elevated levels of butyrate producers may harbor additional beneficial functionalities that influence allergic health outcomes. For example, *Candida albicans*, a yeast commonly found in the gut microbiome, has been shown to promote cross-reactive Th17 cells which trigger an allergic response upon mold exposure,^27^ and *Roseburia* spp., which are butyrate-producers, have been shown to inhibit *C. albicans* growth.^28^ Thus a gut microbiome with higher levels of *Roseburia* may protect against allergic pathology by both producing higher levels of butyrate and by preventing growth of *C. albicans* in the gut. Conversely, the mucin-degrader^29^ *Ruminococcus gnavus* has been shown to precede onset of respiratory allergies and atopic eczema in infants, and *R. gnavus* directly induced a Th2 allergic response and airway hyper-responsiveness in mice.^30^ *R. gnavus* was found to negatively co-occur with many butyrate-producers in multiple independent cohorts. This may be explained by dietary fiber intake, as high dietary fiber promotes the growth of butyrate-producers,^31^ while low dietary fiber promotes the growth of mucin-degraders like *R. gnavus*.^32^ Thus, individuals who consume low fiber diets may have a gut microbiome marked by both lower levels of butyrate producers and higher levels *R. gnavus*, potentially predisposing them to allergic pathology via two synergistic mechanisms: lack of butyrate and *R. gnavus*-driven allergic inflammation. Together, this suggests that microbiome:environment interactions may be driven by synergistic effects of gut microbial ecology, including the presence of beneficial bacteria such as butyrate producers and absence of microbes that promote allergic pathology such as *C. albicans* and *R. gnavus*.

Our cohort was unique in its inclusion of study participants from four distinct neighborhoods in the Houston area, one of the most racially and ethnically diverse metropolitan areas in the United States. While the association between race/ethnicity and the human microbiome is well-established,^33^ we discovered that neighborhood-level geographical associations, including indices of neighborhood-level poverty, are also strongly associated with microbiome composition. While this undoubtedly is partly driven by strong associations between race, ethnicity, neighborhood, and poverty, it reveals these factors should be considered when interpreting reported differences in microbiome composition by racial and ethnic variation. This is particularly apparent when considering the nasal microbiome, as the microbiota detected in the nose were more strongly associated with neighborhood than race/ethnicity. Gut mycobiome composition showed no significant associations with any demographic variable tested, consistent with previous reports that in contrast to the gut bacterial microbiome, which is relatively stable over time, the gut mycobiome is more dynamic.^34^

We did not find associations between hurricane exposures and microbiome composition, which may be explained by several factors. First, it is possible that the human microbiome is relatively resilient in the face of disasters. Alternatively, it may be that Hurricane Harvey caused shifts in microbiome composition that were overpowered by the demographic variation in the microbiome; thus, a more racially and geographically homogenous cohort may have revealed differences not detected in our study. Third, it is possible that the impact of the hurricane on the microbiome is not uniform across individuals and that unique changes occurred in each individual which would not be captured by our methods. As culture-independent sequencing methods cannot profile absolute magnitude of microbes present, it is possible that the amount of microbes (such as mold) significantly varied by hurricane exposures, but were not detected due to this inherent limitation of our methodology.

While we were understandably unable to collect pre-hurricane samples for this study, the lack of association between hurricane exposures and microbiome composition suggests that the robust association between allergic symptoms and the gut microbiome detected in this cohort was likely not driven by hurricane-induced changes in gut microbiota. Rather, the level of butyrate-producing bacteria in each individual’s gut microbiome may have been established prior to the hurricane, and the widespread mold exposure after the hurricane subsequently induced an immune response in susceptible individuals. Furthermore, by collecting self-reported symptoms known to be associated with mold exposure, we were able to capture health outcomes in populations who may have lacked access to health care or were unable to seek medical care for other reasons, a weakness of prior studies assessing health outcomes following flooding events.^6^ However, our study was limited by the restraints inherent in rapidly launching a disaster microbiome study in response to a hurricane, as we were unable to further validate the etiology of symptoms included in our questionnaire.

The lack of association between mycobiome profiles and reported mold exposure suggests that mycobiome sequencing may not adequately capture clinically-significant mold exposures. For the house swabs collected at 1-month post-Harvey only, this may be explained by the use of non-sterile swabs to collect house threshold samples, which were utilized due to a shorthand of supplies immediately after the hurricane, as sterile swabs were prioritized for human sampling. For all other samples types and time points (which utilized sterile collection materials), this lack of association may be explained by several possible factors: 1) the inability of ITS2 sequencing to quantify the magnitude of mold exposure, 2) fungal taxonomy analysis may not be sufficient to delineate the features of mold that trigger an allergic response, as many antigens contain cross-reactive epitopes that are shared among distantly-related fungal and non-fungal species,^35^ 3) the nasal mycobiome may be relatively transient, thus exposures that occurred hours or days prior may not be accurately reflected at the time of nasal sampling. Finally, subjects were only able to swab a small portion of their homes, which assuredly did not capture the full profile of microbes present in the home environment.

Together, our data suggest that the human microbiome is shaped by long-term environmental factors such as neighborhood of residence, race/ethnicity, and socioeconomic status; in addition, it may be relatively resilient and stable in the face of acute, novel microbial exposures from flooding events. We found that exposure to mold is associated with increased risk of allergic symptoms after a severe flooding event. Furthermore, we found that the risk of these allergic symptoms may be dependent on the interaction between the microbiome and the environment, whereby lower levels of microbial butyrate production in the gut may prime individuals for an allergic response to mold, which then manifests upon mold exposure.

## Methods

### Study Design and Recruitment

The Hurricane Harvey Health (Houston3H) Study was launched by Baylor College of Medicine (BCM), The University of Texas Health Science Center (UTHealth), and Oregon State University (OSU) in response to the 2017 hurricane. Leveraging an existing disaster protocol at OSU, rapid IRB approval was obtained at BCM (Protocol H-42111) within days of reopening after the hurricane, and UTHealth obtained reciprocal approval. This study complied with all relevant ethical regulations. Enrollment took place September 23 – October 3, 2017 (referred to as 1-month post-Harvey), with follow-up one year later (August – December 2018; i.e., 12-months post-Harvey). Study eligibility criteria were (1) impacted by Hurricane Harvey through flooding and/or involvement in clean-up efforts, (2) age 5 or older, and (3) conversant in English or Spanish. Enrollment sites in three Harris county neighborhoods highly impacted by flooding were selected for recruitment: Addicks, Baytown, and East Houston. Baylor College of Medicine served as the fourth enrollment site. Written informed consent was obtained in English or Spanish by trained study personnel. Participants at 1-month post-Harvey were re-contacted for follow-up at 12-months post-Harvey; however, participation at 1-month post-Harvey was not a prerequisite for participation at 12-months post-Harvey.

### Exposures and Health Questionnaire

All subjects completed a questionnaire detailing (1) subject demographics and addresses, (2) individual hurricane exposures and clean-up effort involvement, and (3) health outcomes including symptoms associated with mold exposure. Questionnaire data were coded using Research Electronic Data Capture (REDCap) software. After data cleaning, questionnaire data was imported into R (version 3.6.1) for integration with microbiome analyses. Hurricane exposures assessed at both time points included (1) if subjects’ home flooded during Hurricane Harvey, (2) if there were new signs of mold growth in their home after Harvey, (3) if they were rescued during Harvey, and (4) if they were exposed to dirty water, sewage, or visible mold. At 1-month post-Harvey only, the questionnaire additionally asked if subjects were involved in clean-up efforts such as removing mud and debris. Health outcomes assessed included symptoms known to be associated with indoor dampness and/or mold exposure including throat irritation, sinus irritation, eye irritation, wheezing, cough, shortness of breath, and skin rash^7,13^ (1-month and 12-months post-Harvey), and worsening asthma (only assessed at 1-month post-Harvey), collectively described as “allergic symptoms.” For all symptoms except worsening asthma, subjects were asked to report symptoms that had occurred since Hurricane Harvey, excluding symptoms caused by a cold or seasonal allergies, and subjects provided a Yes/No response for each symptom. For worsening asthma, subjects were asked “Has your asthma gotten worse since Hurricane Harvey?” and subjects provided a Yes/No/Don’t Know response. This question was not included in the 12-month post-Harvey questionnaire, and subjects who did not have asthma were instructed to skip this question. Participants at both time points were also asked 1) if they had taken any antibiotics within the past month and/or the past 6 months, 2) if they had taken any probiotics within the past month and/or the past 6 months, and 3) if they are currently or have ever been vegan or vegetarian. For subjects who participated at both time points, answers to similar questions provided at 1-month versus 12-months post-Harvey were at times discrepant; attempts were made to re-contact participants to clarify answers when possible, otherwise data were unchanged.

### Neighborhood Assignment and Area-level Socioeconomic Disadvantage

Subjects’ addresses were geocoded using ArcGIS, and all subjects living within a 7.5 mile radius of the centroid of each study neighborhood were assigned to that neighborhood regardless of their enrollment site. Subjects whose homes were outside the 7.5 mile radius of all neighborhoods were assigned to “Other.” Subject address was also used to assess census tract-level socioeconomic disadvantage by calculating an Area Deprivation Index (ADI) score for each address. Briefly, ADI uses 17 U.S. Census measures of income, housing, employment, and education to measure the level of deprivation within each census tract, with a higher ADI score indicative of greater socioeconomic disadvantage. ADI scores were calculated using the U.S. Census 2012-2016 American Community Survey (ACS) 5-year estimates data summarized to the census tract level. Singh’s formula^36,37^ was used to compute ADI scores for all census tracts in the state of Texas (n = 5,265). Houston3H Study participant addresses were then geocoded using ArcGIS, and participants were assigned an ADI Score based on the ADI score of the census tract they resided in. The median ADI score for the 1-month post-Harvey participants (106.23) was used to stratify all participants into low (ADI Score < 106.23) and high (ADI Score ≥ 106.23) ADI groups. Map of participant locations were created using the R package ggmap using address latitude and longitude geocoded in ArcGIS. For participants who had moved due to Hurricane Harvey, the address of their home at the time of Hurricane Harvey was used for neighborhood assignment and ADI score calculation.

### Sample Collection

Four types of microbiome samples were collected from participants at both time points: nasal, saliva, stool, and swabs of subjects’ homes. Nasal and saliva samples were collected upon enrollment and immediately placed on dry ice. For nasal samples, study personnel swabbed subjects’ nares using the Catch-All Sample Collection Swab (Epicentre) for collection at 1-month post-Harvey and the BBL CultureSwab EZ (BD) at 12-months post-Harvey. Discontinuation of the Catch-All swab prevented the same instrument from being used at both time points. For saliva samples, subjects collected their own unstimulated saliva in a sterile Thermo Nunc 15mL conical tube (Thermo Fisher). At the time of enrollment, subjects were also provided an Omnigene Gut collection kit (DNA Genotek) as well as a swab for at-home collection of stool and a home environment sample, respectively. Due to shorthand of supplies at 1-month post-Harvey, non-sterile Q-tips were provided for house threshold swabs to prioritize sterile collection swabs for human microbiome sampling. At 12-months post-Harvey, a sterile double-tipped BBL Culture Swab (BD) was provided for door threshold sampling. Instructions for stool and home environment collection were given in person and provided in written English and Spanish. Participants were instructed to swab the entry/threshold of their front door. Stool and environmental samples were collected from participants one week after enrollment. A total of 625 samples were collected and processed for subsequent analysis at 1-month post-Harvey (107 house swabs, 120 stool samples, 202 nasal swabs, 196 saliva samples) and 968 samples at 12-months post-Harvey (237 house swabs, 231 stool samples, 255 nasal swabs, and 245 saliva samples).

### Sample Extraction

Microbial DNA was extracted from all sample types using the Qiagen MagAttract PowerSoil DNA Kit. DNA yielded from this extraction was subsequently used for all sequencing.

### 16S rRNA Gene Sequencing

All sample types were profiled by 16S-V4 sequencing. The 16S rRNA V4 region was amplified using 515F and 806R PCR primers containing single-index barcodes and Illumina adapters^38^. Samples were sequenced on the Illumina MiSeq platform using reagent kit v2 (2 × 250 bp) paired-end protocol. Reads were demultiplexed using the Illumina ‘bcl2fastq’ software, then demultiplexed fastq read pairs were merged using USEARCH v7.0.1090^39^ ‘fastq_mergepairs’ function. Merging parameters required read pairs overlap by at least 50 base pairs, a merged length of at least 252 base pairs, a truncation quality above 5, and zero differences in the overlapping region. Merged reads with maximum expected error greater than 0.05 were filtered using usearch70 ‘fastq_filter’ program, and PhiX was filtered using bowtie2 v.2.3.4.3^40^ at the ‘very-sensitive’ parameter setting. Reads were run through usearch70 ‘derep_fulllength’ program and sorted by size using usearch70 ‘sortbysize’ program. Operational Taxonomic Unit (OTU) clustering was performed using the UPARSE algorithm^41^ with a 97% similarity cutoff value. A clustered OTU file with chimeras filtered was created using the usearch70 ‘uchime_ref’ program and the GOLD database.^42,43^ OTUs were mapped against an optimized version of SILVA Database^44^ v132 containing only sequences from the V4 region of the 16S rRNA gene, using usearch70 ‘usearch_global’ function with an identity threshold of 96.8%. All singleton and non-bacterial OTUs were filtered prior to analysis. After filtering, a total of 18,285,084 reads were retained (median 11,629 reads per sample).

### ITS2 Gene Sequencing and Processing

House, nasal, and stool samples were profiled by ITS2 sequencing. ITS3 and ITS4^45^ primers containing adapters for MiSeq sequencing and 12mer molecular barcodes were used to amplify the ITS2 region. Samples were sequenced on the MiSeq platform (Illumina) using the 2×300 bp paired-end protocol^38^. USEARCH v7.0.1090^39^ was used to demultiplex and merge paired reads. Mismatches were allowed for up to 5% of the overlapping sequence; the base with the higher Q score was chosen when there was a mismatch. Reads containing above 0.5% expected errors were discarded. The UPARSE algorithm^41^ was used to iteratively cluster sequences into OTUs at a similarity cutoff value of 99%. USEARCH v8.0.1517 and UCHIME were used to filter chimeras. To identify OTU taxonomy, USEARCH v8.0.1517 was used to map OTUs to the combined GenBank Plant (containing fungi) and Environmental databases. All non-fungal OTUs were filtered prior to analysis. After filtering, a total of 38,140,016 reads were retained (median 33,848 reads per sample).

### Whole Genome Shotgun (WGS) Sequencing and Processing

Stool samples were profiled by WGS sequencing. Metagenomic shotgun sequencing was performed on a NovaSeq 6000 (Illumina) yielding 150 bp paired-end reads. Bbduk3 (BBMap version 38.69) was used for quality trimming and adapter removal from raw fastq reads (trimming parameters: kmer length of 19, one mismatch allowed, and a min Phred quality score of 20). After trimming, reads with a minimum average quality score less than 17 and length shorter than 50 bp were removed. To identify PhiX (standard Illumina spike in) and human (host) reads, bbmap^46^ (version 37.58) was used to map reads to a combined PhiX and hg38 reference database using a kmer length of 15, the bloom filter enabled, and fast search settings, and these reads were subsequently filtered using a custom in-house script. A total of 22,088,152,115 reads were retained after filtering (median 60,319,766 reads per sample, median coverage 9.05 Gbp).Taxonomic profiling was performed using MetaPhlAn2,^47^ and all non-bacterial taxa were filtered prior to analysis. Functional profiling of the gut microbiome was performed using HUMAnN2^48^, with minor modifications to the standard workflow: diamond^49^ (version 0.9.26) was used for filtering and bbmap^46^ was used as the nucleotide aligner. Uniref gene families were then mapped to KEGG Orthologs (KO’s) using the utility mapping file provided by HUMAnN2 (map_ko_unrief90.txt.gz). KO’s were then mapped to KEGG Pathways using the HUMAnN-v0.99^50^ KEGG Pathway database (keggc). Unmapped, unintegrated, and ungrouped results were filtered, and outputs were then normalized to relative abundances for analysis. Butyrate producing-bacteria were determined by searching the UniProt database for bacterial species with an annotated or probable *buk* gene and by searching the literature^51^ for experimentally-verified butyrate production by individual species. Where these results conflicted, experimentally validated butyrate production was used.

### Statistical Analysis

All statistical analysis was performed in R (v3.6.1). Odds ratios and corresponding 95% confidence intervals and p-values were calculated using unconditional logistic regression adjusting for age, sex, race/ethnicity, and education level. Taxonomic analysis was performed using phyloseq (v1.30.0) and vegan (v2.5.6). For all alpha and beta diversity analyses, rarefaction was performed to 5,000 reads (16S), 10,000 reads (ITS2 – nasal and house swabs), 1,000 reads (ITS2 – stool), and 2,982,616 estimated counts (WGS, minimum sample count). House microbiome samples submitted by subjects living in the same house were included in all analyses unless otherwise indicated. All beta diversity analyses used Binary Jaccard distances. For principal coordinate analyses, all samples lacking necessary metadata were removed before analysis, then ordination was performed on Binary Jaccard distance matrices for remaining samples. The adonis function (vegan) was used to calculate R^2^ and a p-values (PERMANOVA), and Bonferroni correction was used to correct for multiple hypothesis testing. PERMDISP was calculated using the betadisper function (vegan) using the group centroid. Alpha diversity was calculated based on the number of unique OTUs (ITS2) or number of unique bacterial species (WGS) detected after rarefaction. Linear Discriminant Analysis Effect Size (LEfSe, online galaxy version)^15^ was used to evaluate bacterial species and pathways that were differentially abundant between subjects with and without allergic symptoms. For taxonomic analysis, relative abundance outputs of MetaPhlAn2 were used as input for LEfSe analysis. For functional pathway analysis, all KEGG pathways with median non-zero coverage > 0.1 were used as input for LEfSe analysis. Co-occurrence analyses were performed with the package co-occur (v1.3) by converting species relative abundance to presence (relative abundance > 0%) or absence (relative abundance = 0%). The R package CuratedMetagenomicData^52^ (v1.16.0) was used to import previously published^33,53–55^ metagenomic data sets for co-occurrence analysis. All statistical tests are indicated in figure legends. Race/Ethnicity was treated as five separate categories (Non-Hispanic Black, Non-Hispanic White, Asian, Hispanic, and Other) for all analyses except for Supplementary Tables 1-3, where Asian was counted as “Other” to protect subject confidentiality. For figures that utilize boxplots, individual data points were plotted using the R function “geom_jitter.”

## Data Availability

16S-V4, ITS2, and WGS metagenomic data will be made available in a publicly accessible repository. Accompanying metadata will be made available, though some variables will be filtered and/or collapsed to maintain subject confidentiality.

## Acknowledgements

Funding for this study included the National Institute of Environmental Health Sciences (NIEHS) Mechanism for Time-Sensitive Research Opportunities in Environmental Health Sciences (R21ES029616 to MB and AO, R21ES029493 to JFP and CLW); BCM Dan L. Duncan Comprehensive Cancer Center, Vivian L. Smith Foundation, 5F30HD090891-03 to KMP, and P30ES030285 to CLW. We are grateful to L. D’Amico, A.N. Quach, and T. Auchtung who helped facilitate enrollment and sample collection.

## Author Contributions

JP, CW, AO, and MB conceived of the study, obtained IRB approval, and launched the study. CS, JS, GA, DNL, MB, KH and CW facilitated subject enrollment and/or sample collection. JS entered and cleaned questionnaire data. AO calculated neighborhood assignments and ADI scores. KH managed microbiome sample processing and sequencing. KP performed all analyses and wrote the manuscript. All authors edited and reviewed the manuscript.

## Code Availability

Analyses were performed using publicly available software.

## References

1. Administration, N. O. and A. Costliest U.S. Tropical Cyclones. https://www.ncdc.noaa.gov/billions/dcmi.pdf (2020).

2. Morast, R. Hurricane Harvey by the numbers. Houston Chronicle https://www.houstonchronicle.com/life/article/Hurricane-Harvey-by-the-numbers-12172287.php (2017).

3. Hunn, D., Dempsey, M. & Zaveri, M. Harvey’s floods. Houston Chronicle https://www.houstonchronicle.com/news/article/In-Harvey-s-deluge-most-damaged-homes-were-12794820.php (2018).

4. Hamblin, J. The Looming Consequences of Breathing Mold. The Atlantic https://www.theatlantic.com/health/archive/2017/08/mold-city/538224/ (2017).

5. Bush, R. K., Portnoy, J. M., Saxon, A., Terr, A. I. & Wood, R. A. The medical effects of mold exposure. J. Allergy Clin. Immunol. 117, 326–333 (2006).

6. Barbeau, D. N., Grimsley, L. F., White, L. E., El-Dahr, J. M. & Lichtveld, M. Mold Exposure and Health Effects Following Hurricanes Katrina and Rita. Annu. Rev. Public Health 31, 165–178 (2010).

7. Mendell, M. J., Mirer, A. G., Cheung, K., Tong, M. & Douwes, J. Respiratory and allergic health effects of dampness, mold, and dampness-related agents: A review of the epidemiologic evidence. Environ. Health Perspect. 119, 748–756 (2011).

8. Chow, N. A. et al. Hurricane-associated mold exposures among patients at risk for invasive mold infections after hurricane harvey — Houston, Texas, 2017. Morb. Mortal. Wkly. Rep. 68, 470–473 (2019).

9. Pascal, M. et al. Microbiome and allergic diseases. Front. Immunol. 9, 1584 (2018).

10. Liu, H. et al. Butyrate: A double-edged sword for health? Adv. Nutr. 9, 21–29 (2018).

11. Theiler, A. et al. Butyrate ameliorates allergic airway inflammation by limiting eosinophil trafficking and survival. J. Allergy Clin. Immunol. 144, 764–776 (2019).

12. Oluyomi, A. O. et al. Houston hurricane Harvey health (Houston-3H) study: assessment of allergic symptoms and stress after hurricane Harvey flooding. Environ. Heal. A Glob. Access Sci. Source 20, 1–15 (2021).

13. Mold. https://www.cdc.gov/mold/ (2019).

14. Fujimura, K. E. & Lynch, S. V. Microbiota in allergy and asthma and the emerging relationship with the gut microbiome. Cell Host Microbe 17, 592–602 (2015).

15. Segata, N. et al. Metagenomic biomarker discovery and explanation. Genome Biol. 12, R60 (2011).

16. Corrêa-Oliveira, R., Fachi, J. L., Vieira, A., Sato, F. T. & Vinolo, M. A. R. Regulation of immune cell function by short-chain fatty acids. Clin. Transl. Immunol. 5, 1–8 (2016).

17. Turnbaugh, P. J. et al. An obesity-associated gut microbiome with increased capacity for energy harvest. Nature 444, 1027–1031 (2006).

18. Francino, M. P. Antibiotics and the human gut microbiome: Dysbioses and accumulation of resistances. Front. Microbiol. 6, 1–11 (2016).

19. Nylund, L. et al. Severity of atopic disease inversely correlates with intestinal microbiota diversity and butyrate-producing bacteria. Allergy Eur. J. Allergy Clin. Immunol. 70, 241–244 (2015).

20. Roduit, C. et al. High levels of butyrate and propionate in early life are associated with protection against atopy. Allergy Eur. J. Allergy Clin. Immunol. 74, 799–809 (2019).

21. Cait, A. et al. Reduced genetic potential for butyrate fermentation in the gut microbiome of infants who develop allergic sensitization. J. Allergy Clin. Immunol. 144, 1638-1647.e3 (2019).

22. Wang, Q. et al. A metagenome-wide association study of gut microbiota in asthma in UK adults. BMC Microbiol. 18, 1–7 (2018).

23. Trompette, A. et al. Gut microbiota metabolism of dietary fiber influences allergic airway disease and hematopoiesis. Nat. Med. 20, 159–166 (2014).

24. Vieira, R. de S. et al. Butyrate Attenuates Lung Inflammation by Negatively Modulating Th9 Cells. Front. Immunol. 10, 67 (2019).

25. Halnes, I. et al. Soluble fibre meal challenge reduces airway inflammation and expression of GPR43 and GPR41 in asthma. Nutrients 9, (2017).

26. McLoughlin, R. et al. Soluble fibre supplementation with and without a probiotic in adults with asthma: A 7-day randomised, double blind, three way cross-over trial. EBioMedicine 46, 473–485 (2019).

27. Bacher, P. et al. Human Anti-fungal Th17 Immunity and Pathology Rely on Cross-Reactivity against Candida albicans. Cell 176, 1–16 (2019).

28. García, C. et al. The Human Gut Microbial Metabolome Modulates Fungal Growth via the TOR Signaling Pathway. mSphere 2, 1–15 (2017).

29. Crost, E. H. et al. The mucin-degradation strategy of Ruminococcus gnavus: The importance of intramolecular trans-sialidases. Gut Microbes 7, 302–312 (2016).

30. Chua, H. H. et al. Intestinal Dysbiosis Featuring Abundance of Ruminococcus gnavus Associates With Allergic Diseases in Infants. Gastroenterology 154, 154–167 (2018).

31. Tomova, A. et al. The effects of vegetarian and vegan diets on gut microbiota. Front. Nutr. 6, (2019).

32. Desai, M. S. et al. A Dietary Fiber-Deprived Gut Microbiota Degrades the Colonic Mucus Barrier and Enhances Pathogen Susceptibility. Cell 167, 1339-1353.e21 (2016).

33. Huttenhower, C. et al. Structure, function and diversity of the healthy human microbiome. Nature 486, 207–214 (2012).

34. Nash, A. K. et al. The gut mycobiome of the Human Microbiome Project healthy cohort. Microbiome 5, 153 (2017).

35. Ursula, B. S., Verena, D., Raphaela, P. & Breitenbach, M. The Spectrum of Fungal Allergy. 58–86 (2008). doi:10.1159/000107578

36. Knighton, A. J., Savitz, L., Belnap, T., Stephenson, B. & VanDerslice, J. Introduction of an Area Deprivation Index Measuring Patient Socio-economic Status in an Integrated Health System: Implications for Population Health. eGEMs (Generating Evid. Methods to Improv. patient outcomes) 4, 9 (2016).

37. Singh, G. K. Area Deprivation and Widening Inequalities in US Mortality, 1969-1998. Am. J. Public Health 93, 1137–1143 (2003).

38. Caporaso, J. G. et al. Ultra-high-throughput microbial community analysis on the Illumina HiSeq and MiSeq platforms. ISME J. 6, 1621–1624 (2012).

39. Edgar, R. C. Search and clustering orders of magnitude faster than BLAST. Bioinformatics 26, 2460–2461 (2010).

40. Langmead, B. & Salzberg, S. L. Fast gapped-read alignment with Bowtie 2. Nat. Methods 9, 357–359 (2012).

41. Edgar, R. C. UPARSE: Highly accurate OTU sequences from microbial amplicon reads. Nat. Methods 10, 996–998 (2013).

42. Kyrpides, N. C. Genomes OnLine Database (GOLD 1.0): a monitor of complete and ongoing genome projects world-wide. Bioinformattics 15, 773–774 (1999).

43. Mukherjee, S. et al. Genomes OnLine database (GOLD) v.7: Updates and new features. Nucleic Acids Res. 47, D649–D659 (2019).

44. Quast, C. et al. The SILVA ribosomal RNA gene database project: Improved data processing and web-based tools. Nucleic Acids Res. 41, 590–596 (2013).

45. White, T., Bruns, T., Lee, S. & Taylor, J. Amplification and direct sequencing of fungal ribosomal RNA genes for phylogenetics. In PCR Protocols: A Guide to Methods and Applications. (1990).

46. Bushnell, B. BBMap. sourceforge.net/projects/bbmap/

47. Segata, N. et al. Metagenomic microbial community profiling using unique clade-specific marker genes. Nat. Methods 9, 811–814 (2012).

48. Franzosa, E. A. et al. Species-level functional profiling of metagenomes and metatranscriptomes. Nat. Methods 15, 962–968 (2018).

49. Buchfink, B., Xie, C. & Huson, D. H. Fast and sensitive protein alignment using DIAMOND. Nat. Methods 12, 59–60 (2014).

50. Abubucker, S. et al. Metabolic reconstruction for metagenomic data and its application to the human microbiome. PLoS Comput. Biol. 8, e1002358 (2012).

51. Louis, P. & Flint, H. J. Formation of propionate and butyrate by the human colonic microbiota. Environ. Microbiol. 19, 29–41 (2017).

52. Pasolli, E. et al. Accessible, curated metagenomic data through ExperimentHub. Nat. Methods 14, 1023–1024 (2017).

53. Schirmer, M. et al. Linking the Human Gut Microbiome to Inflammatory Cytokine Production Capacity. Cell 167, 1125-1136.e8 (2016).

54. Le Chatelier, E. et al. Richness of human gut microbiome correlates with metabolic markers. Nature 500, 541–6 (2013).

55. Dhakan, D. B. et al. The unique composition of Indian gut microbiome, gene catalogue, and associated fecal metabolome deciphered using multi-omics approaches. Gigascience 8, 1–20 (2019).

