## Supplemental Tables and Figures for "Butyrate-producing gut bacteria are associated with protection from allergic symptoms after Hurricane Harvey"

**Supplementary Table 1.** Characteristics of Houston3H subjects across Houston-area neighborhoods.

| Characteristic | 1-month post-Harvey | | | | | 12-months post-Harvey | | | | |
|---|---|---|---|---|---|---|---|---|---|---|
| | Addicks **N (%)** | Baytown **N (%)** | Bellaire-Meyerland **N (%)** | East Houston **N (%)** | Other **N (%)** | Addicks **N (%)** | Baytown **N (%)** | Bellaire-Meyerland **N (%)** | East Houston **N (%)** | Other **N (%)** |
| **Race/Ethnicity** | | | | | | | | | | |
| Black (NH) | 2 (7.1%) | 49 (81.7%) | 0 (0%) | 22 (42.3%) | 10 (41.7%) | 0 (0%) | 34 (77.3%) | 1 (2.1%) | 44 (53.7%) | 8 (22.2%) |
| White (NH) | 21 (75.0%) | 3 (5.0%) | 30 (73.2%) | 0 (0%) | 6 (25.0%) | 50 (89.3%) | 2 (4.5%) | 35 (72.9%) | 3 (3.7%) | 16 (44.4%) |
| Hispanic | 3 (10.7%) | 2 (3.3%) | 1 (2.4%) | 30 (57.7%) | 4 (16.7%) | 5 (8.9%) | 2 (4.5%) | 3 (6.2%) | 34 (41.5%) | 6 (16.7%) |
| Other[a] | 2 (7.1%) | 6 (10.0%) | 10 (24.4%) | 0 (0%) | 4 (16.7%) | 1 (1.8%) | 6 (13.6%) | 9 (18.8%) | 1 (1.2%) | 6 (16.7%) |
| Missing | 0 | 0 | 0 | 1 | 0 | 0 | 0 | 0 | 0 | 0 |
| **Area Deprivation Index (ADI)** | | | | | | | | | | |
| High ADI | 2 (7.1%) | 42 (70.0%) | 3 (7.3%) | 52 (98.1%) | 5 (20.8%) | 0 (0%) | 33 (75%) | 2 (4.2%) | 81 (98.8%) | 7 (19.4%) |
| Low ADI | 26 (92.9%) | 18 (30.0%) | 38 (92.7%) | 1 (1.9%) | 19 (79.2%) | 56 (100%) | 11 (25%) | 46 (95.8%) | 1 (1.2%) | 29 (80.6%) |
| **Age** | | | | | | | | | | |
| Child (<18) | 0 (0%) | 9 (15.0%) | 0 (0%) | 7 (13.2%) | 1 (4.2%) | 1 (1.8%) | 6 (14.0%) | 0 (0%) | 16 (20.0%) | 2 (5.7%) |
| Adult (18-65) | 18 (64.3%) | 31 (51.7%) | 35 (85.4%) | 39 (73.6%) | 22 (91.7%) | 27 (49.1%) | 19 (44.2%) | 39 (81.2%) | 49 (61.3%) | 31 (88.6%) |
| Senior (> 65) | 10 (35.7%) | 20 (33.3%) | 6 (14.6%) | 7 (13.2%) | 1 (4.2%) | 27 (49.1%) | 18 (41.9%) | 9 (18.8%) | 15 (18.8%) | 2 (5.7%) |
| Missing | | | | | | 1 | 1 | 0 | 2 | 1 |
| **Sex** | | | | | | | | | | |
| Female | 18 (64.3%) | 42 (70.0%) | 29 (70.7%) | 41 (77.4%) | 18 (75.0%) | 32 (57.1%) | 30 (68.2%) | 32 (66.7%) | 53 (64.6%) | 27 (75.0%) |
| Male | 10 (35.7%) | 18 (30.0%) | 12 (29.3%) | 12 (22.6%) | 6 (25.0%) | 24 (42.9%) | 14 (31.8%) | 16 (33.3%) | 29 (35.4%) | 9 (25.0%) |

[a]Includes Asian

**Supplementary Table 2.** Characteristics of subjects with and without allergic symptoms, 1-month post-Harvey.

| Characteristic | All Subjects | | | | Subjects with Stool Samples (WGS) | | | |
|---|---|---|---|---|---|---|---|---|
| | No Symptoms (N = 36) **N (%)** | At Least One Symptom (N = 145) **N (%)** | Did not answer (N = 25) **N** | p-value[a] | No Symptoms (N = 18) **N (%)** | At Least One Symptom (N = 88) **N (%)** | Did not answer (N= 11) **N** | p-value[a] |
| **Neighborhood (during Harvey)** | | | | | | | | |
| Addicks | 2 (5.6%) | 24 (16.6%) | 2 | | 1 (5.6%) | 20 (22.7%) | 1 | |
| Baytown | 9 (25.0%) | 46 (31.7%) | 5 | 0.054[b] | 3 (16.7%) | 21 (23.9%) | 0 | 0.17[b] |
| Bellaire/Meyerland | 8 (22.2%) | 33 (22.8%) | 0 | | 7 (38.9%) | 24 (27.3%) | 0 | |
| East Houston | 14 (38.9%) | 24 (16.6%) | 15 | | 6 (33.3%) | 13 (14.8) | 8 | |
| Other | 3 (8.3%) | 18 (12.4%) | 3 | | 1 (5.6%) | 10 (11.4%) | 2 | |
| **Race/Ethnicity** | | | | | | | | |
| Black (Non-Hispanic) | 16 (44.4%) | 57 (39.6%) | 10 | | 4 (22.2%) | 26 (29.5%) | 2 | |
| White (Non-Hispanic) | 7 (19.4%) | 52 (36.1%) | 1 | 0.15[b] | 5 (27.8%) | 37 (42.0%) | 1 | 0.13[b] |
| Hispanic | 9 (25.0) | 19 (13.2%) | 12 | | 7 (38.9%) | 12 (13.6%) | 7 | |
| Other[c] | 4 (11.1%) | 16 (11.1%) | 2 | | 2 (11.1%) | 13 (14.8%) | 1 | |
| Missing | 0 | 1 | 0 | | 0 | 0 | 0 | |
| **Area Deprivation Index (ADI)** | | | | | | | | |
| High ADI (lower SES) | 22 (61.1%) | 61 (42.1%) | 21 | 0.06[b] | 8 (44.4%) | 31 (35.2%) | 9 | 0.59[b] |
| Low ADI (higher SES) | 14 (38.9%) | 84 (57.9%) | 4 | | 10 (55.6%) | 57 (64.8%) | 2 | |
| **Age** | | | | | | | | |
| Median | 47.5 | 52.0 | 39.0 | 0.076[d] | 46.5 | 57.0 | 38.0 | 0.008[d] |
| [IQR] | [29.5,56.3] | [39.0,64.0] | [25.0,61.0] | | [30.5, 53.8] | [44.0, 65.0] | [23.5, 51.0] | |
| **Sex** | | | | | | | | |
| Female | 24 (66.7%) | 106 (73.1%) | 18 | 0.53[b] | 10 (55.6%) | 67 (76.1%) | 9 | 0.09[b] |
| Male | 12 (33.3%) | 39 (26.9%) | 7 | | 8 (44.4%) | 21 (23.9%) | 2 | |
| **Education** | | | | | | | | |
| High School or Less | 12 (36.4%) | 27 (19.0%) | 15 | | 5 (31.2%) | 14 (16.1%) | 8 | |
| Some College | 2 (6.1%) | 25 (17.6%) | 5 | 0.10[b] | 2 (12.5%) | 13 (14.9%) | 0 | 0.46[b] |
| Undergraduate Degree | 9 (27.3%) | 50 (35.2%) | 3 | | 3 (18.8%) | 29 (33.3%) | 1 | |
| Advanced Degree | 10 (30.3%) | 40 (28.2%) | 1 | | 6 (37.5%) | 31 (35.6%) | 1 | |
| Missing | 3 | 3 | 1 | | 2 | 1 | 1 | |
| **BMI[e]** | | | | | | | | |
| Underweight (< 18.5) | 0 (0.0%) | 2 (2.6%) | 0 | | 0 (0.0%) | 1 (1.8%) | 0 | |
| Normal weight (18.5-25) | 4 (26.7%) | 29 (38.2%) | 1 | 0.65[b] | 3 (33.3%) | 17 (30.9%) | 1 | 0.45[b] |
| Overweight (25-30) | 7 (46.7%) | 23 (30.3%) | 1 | | 5 (55.6%) | 18 (32.7%) | 1 | |
| Obese (> 30) | 4 (26.7%) | 22 (28.9%) | 1 | | 1 (11.1%) | 19 (34.5%) | 1 | |
| Missing | 21 | 69 | 22 | | 9 | 33 | 8 | |
| **House Flooded** | | | | | | | | |
| No | 8 (22.2%) | 28 (19.3%) | 7 | 0.65[b] | 1 (5.6%) | 13 (14.8%) | 2 | 0.46[b] |
| Yes | 28 (77.8%) | 117 (80.7%) | 18 | | 17 (94.4%) | 75 (85.2%) | 9 | |
| **Asthma** | | | | | | | | |
| No | 30 (88.2%) | 112 (84.8%) | 17 | 0.79[b] | 16 (94.1%) | 69 (85.2%) | 6 | 0.46[b] |
| Yes | 4 (11.8%) | 20 (15.2%) | 0 | | 1 (5.9%) | 12 (14.8%) | 0 | |
| Don't Know/Missing | 2 | 13 | 8 | | 1 | 7 | 5 | |
| **Currently Smoke** | | | | | | | | |
| No | 32 (91.4%) | 128 (89.5%) | 14 | 1[b] | 15 (88.2%) | 81 (93.1%) | 6 | 0.61[b] |
| Yes | 3 (8.6%) | 15 (10.5%) | 3 | | 2 (11.8%) | 6 (6.9%) | 0 | |
| Missing | 1 | 2 | 8 | | 1 | 1 | 5 | |
| **Household member who smokes** | | | | | | | | |
| No | 33 (97.1%) | 127 (90.1%) | 16 | 0.31[b] | 16 (100%) | 80 (94.1%) | 7 | 1[b] |
| Yes | 1 (2.9%) | 14 (9.9%) | 1 | | 0 (0.0%) | 5 (5.9%) | 0 | |
| Missing | 2 | 4 | 8 | | 2 | 3 | 4 | |
| **Antibiotics in past month** | | | | | | | | |
| No | 30 (93.8%) | 119 (86.9%) | 16 | | 16 (100%) | 77 (89.5%) | 5 | |
| Yes | 2 (6.2%) | 18 (13.1%) | 1 | 0.37[b] | 0 (0.0%) | 9 (10.5) | 1 | 0.35[b] |
| Don't Know/Missing | 4 | 8 | 8 | | 2 | 2 | 5 | |
| **Antibiotics in past 6 mo.** | | | | | | | | |
| No | 27 (84.4%) | 109 (78.4%) | 15 | | 16 (100%) | 68 (79.1%) | 4 | |
| Yes | 5 (15.6%) | 30 (21.6%) | 2 | 0.63[b] | 0 (0.0%) | 18 (20.9%) | 2 | 0.07[b] |
| Don't Know/Missing | 4 | 6 | 8 | | 2 | 2 | 5 | |
| **Probiotics in past 6 mo.** | | | | | | | | |
| No | 27 (93.1%) | 102 (76.1%) | 14 | | 13 (86.7%) | 61 (74.4%) | 5 | |
| Yes | 2 (6.9%) | 32 (23.9%) | 1 | 0.045[b] | 2 (13.3) | 21 (25.6%) | 0 | 0.51[b] |
| Don't Know/ Missing | 7 | 11 | 10 | | 3 | 6 | 6 | |
| **Vegetarian or Vegan (Past or Current)** | | | | | | | | |
| No | 32 (100%) | 127 (88.8%) | 16 | 0.046[b] | 16 (100%) | 74 (85.1%) | 6 | 0.21[b] |
| Yes | 0 (0%) | 16 (11.2%) | 0 | | 0 (0.0%) | 13 (14.9%) | 0 | |
| Don't Know/Missing | 4 | 2 | 9 | | 2 | 1 | 5 | |

[a]Does not include subjects who did not answer allergic symptoms questions.
[b]Fisher's Exact Test
[c]Includes Asian
[d]Two-sided Mann-Whitney Test
[e]Height was not assessed at 1-month post-Harvey; calculated BMI for adult (age > 18) longitudinal subjects only using height provided at 12-months post-Harvey and weight provided at 1-month post-Harvey.

**Supplementary Table 3.** Characteristics of subjects with and without allergic symptoms, 12-months post-Harvey.

| **Characteristic** | **All Subjects** | | | | **Subjects with Stool Samples (WGS) With Mold in Home** | | | |
|---|---|---|---|---|---|---|---|---|
| | No Symptoms (N = 82) N (%) | At Least One Symptom (N = 173) N (%) | Did not answer (N = 11) N | p-value[a] | No Symptoms (N = 29) | At Least One Symptom (N = 97) | Did not answer (N = 4) | p-value[a] |
| **Neighborhood (during Harvey)** | | | | | | | | |
| Addicks | 12 (14.6%) | 44 (25.4%) | 0 | | 5 (17.2%) | 28 (28.9%) | 0 | |
| Baytown | 16 (19.5%) | 25 (14.5%) | 3 | 0.34[b] | 3 (10.3%) | 12 (12.4%) | 2 | 0.27[b] |
| Bellaire/Meyerland | 17 (20.7%) | 29 (16.8%) | 2 | | 9 (31.0%) | 13 (13.4%) | 1 | |
| East Houston | 26 (31.7%) | 52 (30.1%) | 4 | | 10 (34.5%) | 33 (34.0%) | 1 | |
| Other | 11 (13.4%) | 23 (13.3%) | 2 | | 2 (6.9%) | 11 (11.3%) | 0 | |
| **Race/Ethnicity** | | | | | | | | |
| Black (Non-Hispanic) | 26 (31.7%) | 54 (31.2%) | 7 | | 10 (34.5%) | 28 (28.9%) | 3 | |
| White (Non-Hispanic) | 30 (36.6%) | 73 (42.2%) | 3 | 0.74[b] | 11 (37.9%) | 42 (43.3%) | 1 | 0.60[b] |
| Hispanic | 19 (23.2%) | 31 (17.9) | 0 | | 7 (24.1%) | 17 (17.5%) | 0 | |
| Other[c] | 7 (8.5%) | 15 (8.7%) | 1 | | 1 (3.4%) | 10 (10.3%) | 0 | |
| **Area Deprivation Index (ADI)** | | | | | | | | |
| High ADI (lower SES) | 41 (50.0%) | 76 (43.9%) | 6 | 0.42[b] | 15 (51.7%) | 43 (44.3%) | 2 | 0.53[b] |
| Low ADI (higher SES) | 41 (50.0%) | 97 (56.1%) | 5 | | 14 (48.3%) | 54 (55.7%) | 2 | |
| **Age** | | | | | | | | |
| Median | 46.5 | 58.0 | 50.0 | 0.005[d] | 46.0 | 58.0 | 57.5 | 0.19[d] |
| [IQR] | [29.0,63.0] | [39.0, 66.0] | [48.0, 65.0] | | [35.0, 60.8] | [39.0, 66.0] | [40.3, 68.5] | |
| Missing | 2 | 1 | 2 | | 1 | 1 | | |
| **Sex** | | | | | | | | |
| Female | 46 (56.1%) | 120 (69.4%) | 8 | 0.05[b] | 13 (48.8%) | 67 (69.1%) | 2 | 0.03[b] |
| Male | 36 (43.9%) | 53 (30.6%) | 3 | | 16 (55.2%) | 30 (30.9%) | 2 | |
| **Education** | | | | | | | | |
| High School or Less | 28 (35.0%) | 45 (26.2%) | 2 | | 9 (32.1%) | 28 (29.2%) | 1 | |
| Some College | 9 (11.2%) | 21 (12.2%) | 1 | 0.42[b] | 4 (14.3%) | 12 (12.5%) | 1 | |
| Undergraduate Degree | 18 (22.5%) | 53 (30.8%) | 3 | | 6 (21.4%) | 27 (28.1%) | 0 | 0.92[b] |
| Advanced Degree | 25 (31.2%) | 53 (30.8%) | 3 | | 9 (32.1%) | 29 (30.2%) | 2 | |
| Missing | 2 | 1 | 2 | | 1 | 1 | 0 | |
| **BMI** | | | | | | | | |
| Underweight (< 18.5) | 3 (3.8%) | 4 (2.5%) | 0 | | 2 (6.9%) | 2 (2.3%) | 0 | |
| Normal weight (18.5-25) | 26 (32.9%) | 58 (36.7%) | 1 | 0.49[b] | 9 (31.0%) | 33 (37.9%) | 1 | 0.57[b] |
| Overweight (25-30) | 26 (32.9%) | 39 (24.7%) | 1 | | 8 (27.6%) | 21 (24.1%) | 1 | |
| Obese (> 30) | 24 (30.4%) | 57 (36.1%) | 0 | | 10 (34.5%) | 31 (35.6%) | 0 | |
| Missing | 3 | 15 | 9 | | 0 | 10 | 2 | |
| **House Flooded** | | | | | | | | |
| No | 15 (18.3%) | 15 (8.7%) | 4 | 0.04[b] | 2 (6.9%) | 3 (3.1%) | 2 | 0.32[b] |
| Yes | 67 (81.7%) | 158 (91.3%) | 6 | | 27 (93.1%) | 94 (96.9%) | 2 | |
| Missing | 0 | 0 | 1 | | | | | |
| **Asthma** | | | | | | | | |
| No | 78 (95.1%) | 151 (87.3) | 11 | 0.07[b] | 27 (93.1%) | 85 (87.6%) | 4 | 0.52[b] |
| Yes | 4 (4.9%) | 22 (12.7%) | 0 | | 2 (6.9%) | 12 (12.4%) | 0 | |
| **Currently Smoke** | | | | | | | | |
| No | 79 (96.3%) | 148 (86.5%) | 3 | 0.02[b] | 28 (96.6%) | 82 (84.5%) | 3 | 0.12[b] |
| Yes | 3 (3.7%) | 23 (13.5%) | 0 | | 1 (3.4%) | 15 (15.5%) | 0 | |
| Missing | 0 | 2 | 8 | | 0 | 0 | 1 | |
| **Household member who smokes** | | | | | | | | |
| No | 76 (93.8%) | 148 (88.1%) | 1 | 0.18[b] | 25 (89.3%) | 82 (86.3%) | 1 | 1[b] |
| Yes | 5 (6.2%) | 20 (11.9%) | 0 | | 3 (10.7%) | 13 (13.7%) | 0 | |
| Missing | 1 | 5 | 10 | | 1 | 2 | 3 | |
| **Antibiotics in past 6 months** | | | | | | | | |
| No | 63 (82.9%) | 98 (62.0%) | 2 | 0.001[b] | 25 (96.2%) | 54 (60.7%) | 2 | 0.0003[b] |
| Yes | 13 (17.1%) | 60 (38.0%) | 0 | | 1 (3.8%) | 35 (39.3%) | 0 | |
| Don't Know/Missing | 6 | 15 | 9 | | 3 | 8 | 2 | |
| **Probiotics in past 6 months** | | | | | | | | |
| No | 62 (82.7%) | 117 (77.0%) | 2 | | 23 (88.5%) | 71 (82.6%) | 2 | |
| Yes | 13 (17.3%) | 35 (23.0%) | 0 | 0.39[b] | 3 (11.5%) | 15 (17.4%) | 0 | 0.56[b] |
| Don't Know/ Missing | 7 | 21 | 9 | | 3 | 11 | 2 | |
| **Vegetarian or Vegan** | | | | | | | | |
| No | 76 (96.2%) | 141 (89.8%) | 2 | 0.13[b] | 26 (96.3%) | 78 (87.6%) | 2 | 0.29[b] |
| Yes | 3 (3.8%) | 16 (10.2%) | 0 | | 1 (3.7%) | 11 (12.4%) | 0 | |
| Don't Know/Missing | 3 | 16 | 9 | | 2 | 8 | 2 | |

[a]Does not include subjects who did not answer allergic symptoms questions.
[b]Fisher's Exact Test
[c]Includes Asian
[d]Two-sided Mann-Whitney test

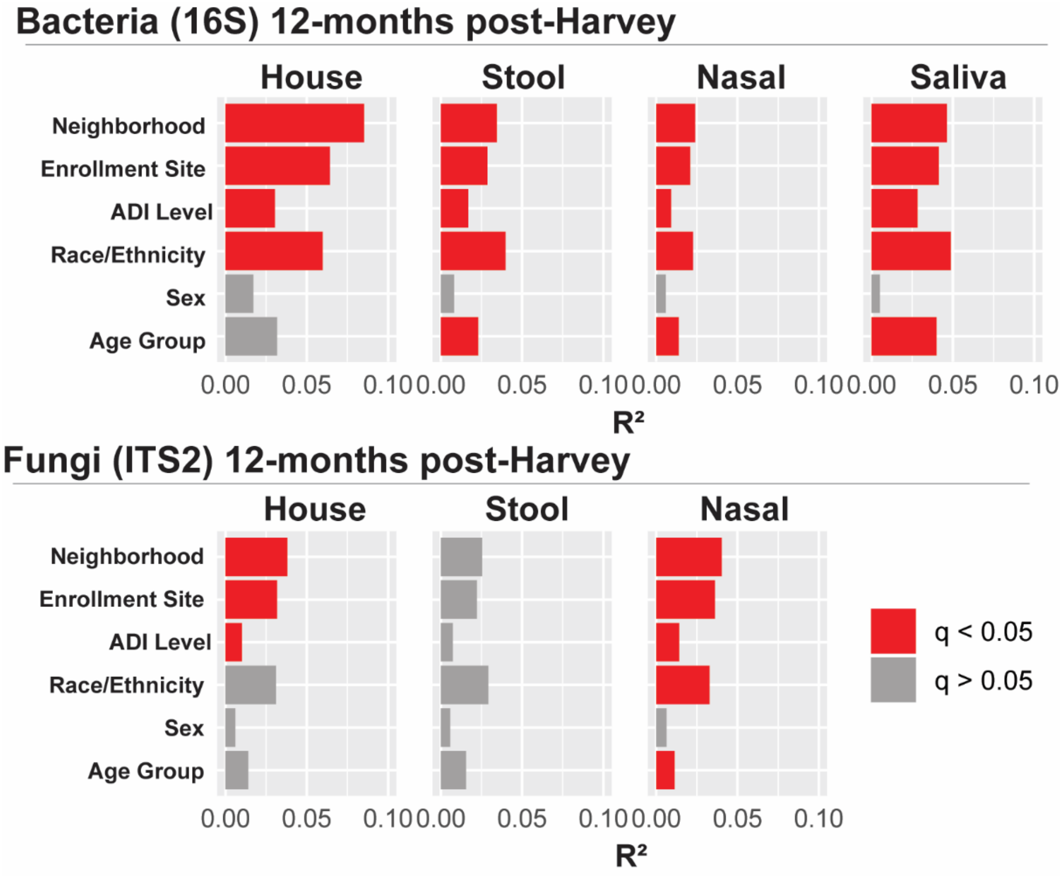


**Supplementary Figure 1. Impact of subject demographics on bacterial and fungal microbiome composition at 12-months post-Harvey.** Principal coordinate analysis (PCoA) of Binary Jaccard distances was used to evaluate associations between demographic variables and microbiome composition at 12-months post-Harvey. PERMANOVA was used to calculate $R^2$ (percent of total microbial variation accounted for by each variable) and p-values. Bonferroni correction of p-values was used to calculate q-values for each body site. Total samples (n) remaining in each analysis after rarefaction: house 16S (n=69), nasal 16S (n=241), stool 16S (n=195), saliva 16S (n=229), house ITS2 (n=147), nasal ITS2 (n=221), stool ITS2 (n=168). House microbiome analysis only includes samples from subjects still living in the same home as they were at the time of Hurricane Harvey and only includes one swab per household. Sample numbers and statistics for each individual analysis are provided in Supplementary Table 4.

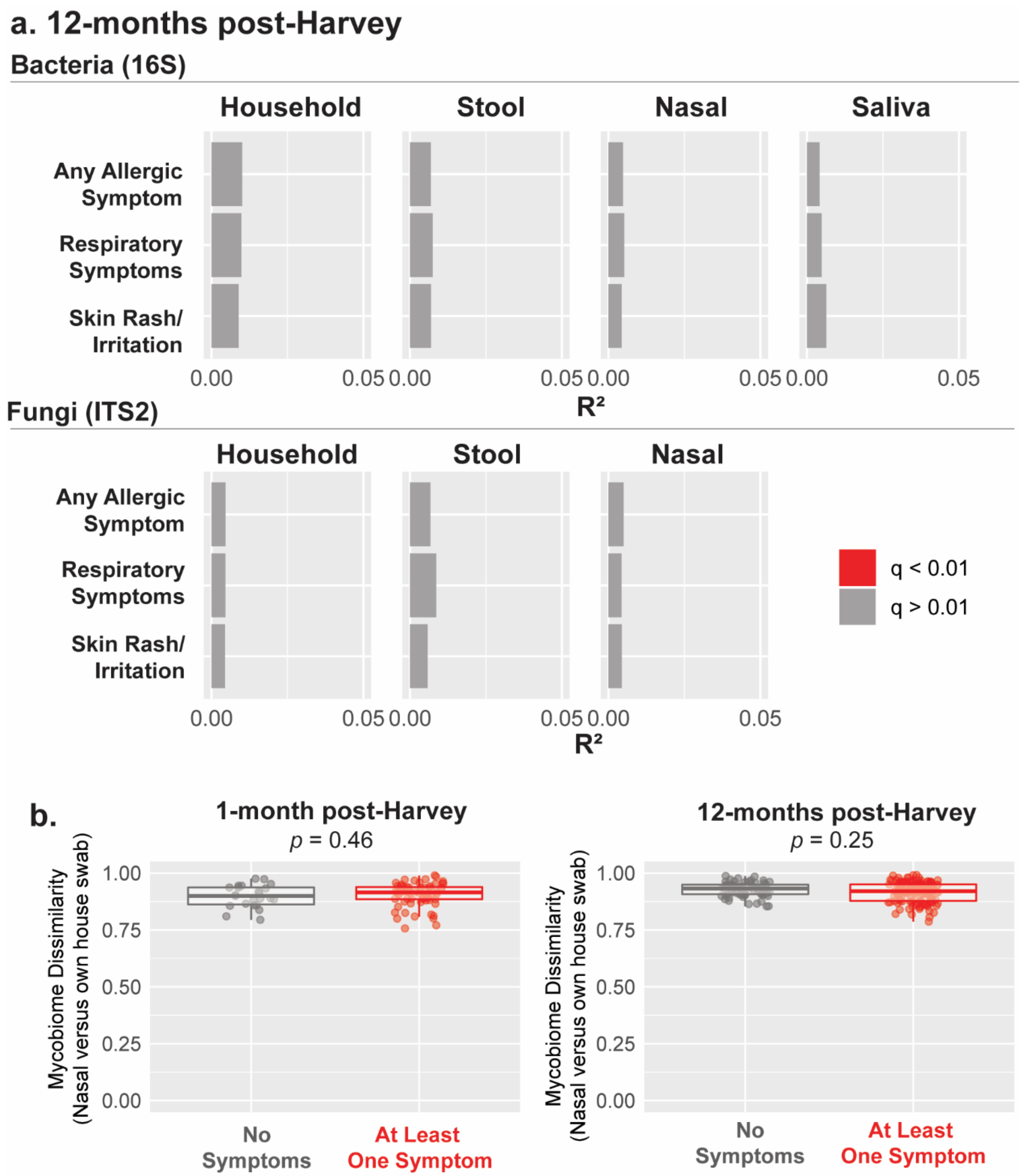


**Supplementary Figure 2. Association between microbiome composition and allergic health outcomes. a.** PCoA analyses of Binary Jaccard distances were used to evaluate associations between bacterial (16S) and fungal (ITS2) microbiota composition and reported allergic symptoms within 12-months post-Harvey. PERMANOVA was used to calculate $R^2$ (percent of total microbial variation accounted for by each variable) and p-values, followed by Bonferroni correction. Total samples in each analysis after rarefaction: house 16S (n=100), stool 16S (n=188), nasal 16S (n=231), saliva 16S (n=219), house ITS2 (n=216), stool ITS2 (n=161), nasal ITS2 (n=213). Sample numbers and statistics for each individual analysis are provided in Supplementary Table 4. **b.** Mycobiome (ITS2) dissimilarity (Binary Jaccard distance) comparing each subject's nasal sample to the swab of their own home reveals allergic symptoms are not associated with higher similarity between the nasal and household mycobiome at 1-month post-Harvey (left, n=23 without symptoms, n=65 with symptoms) and 12-months post-Harvey (right, n=61 without symptoms, n=134 with symptoms) (two-sided Mann-Whitney, $p > 0.05$). Box plots indicate median and interquartile range (IQR), whiskers show smallest (lower whisker) or largest (upper whisker) value within 1.5 times the IQR.

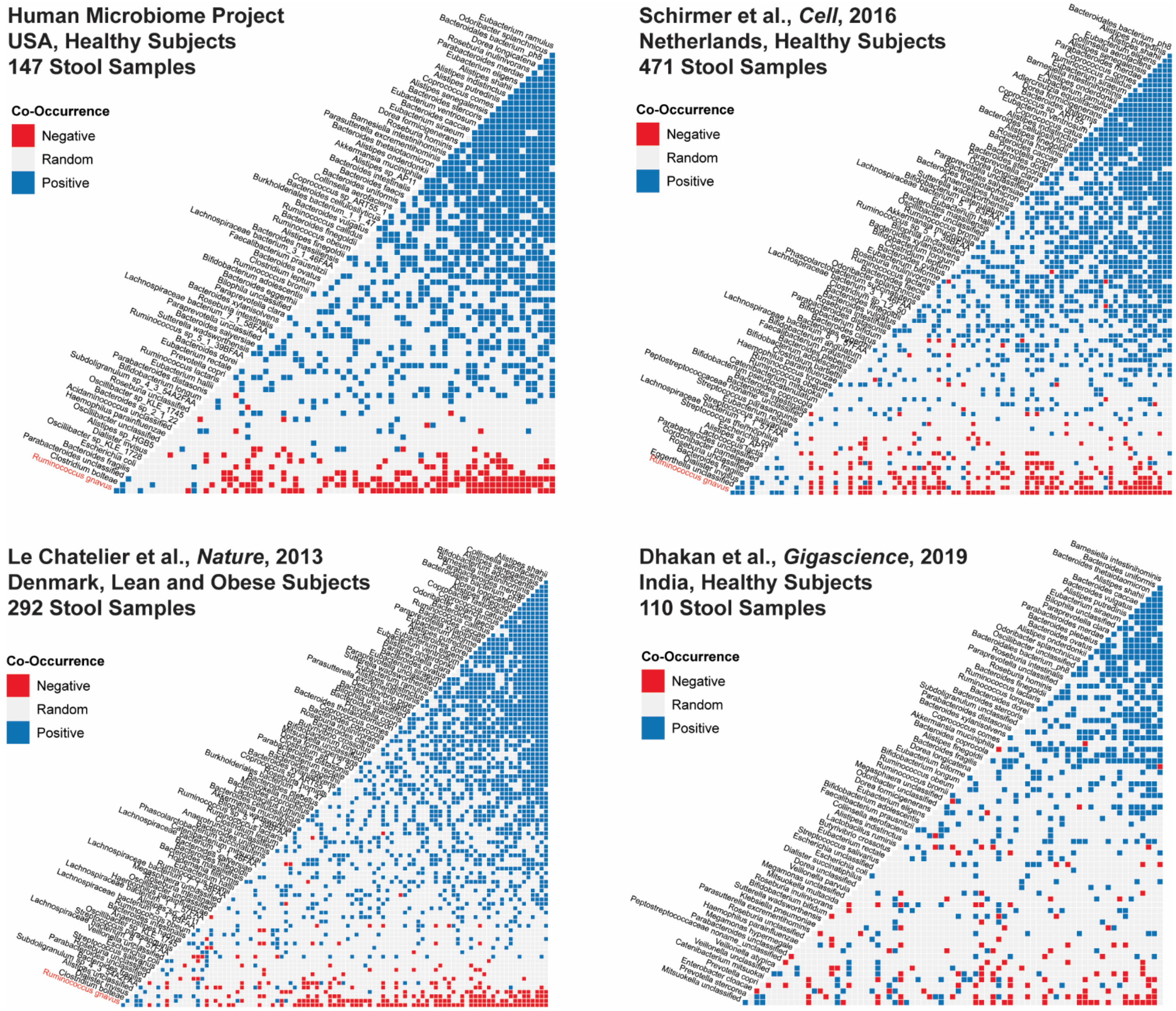


**Supplementary Figure 3. Co-occurrence analysis of gut bacterial species from publicly available metagenomic datasets.** Co-occurrence analysis based on presence/absence of bacterial species reveals positive (blue) and negative (red) associations between bacterial species in previously published cohorts from the US, Europe, and India. Co-occurrence analyses include species present at >0.1% relative abundance in at least 10% of samples. Human Microbiome Project includes 147 samples from 95 subjects, all other cohorts contain samples from unique subjects.

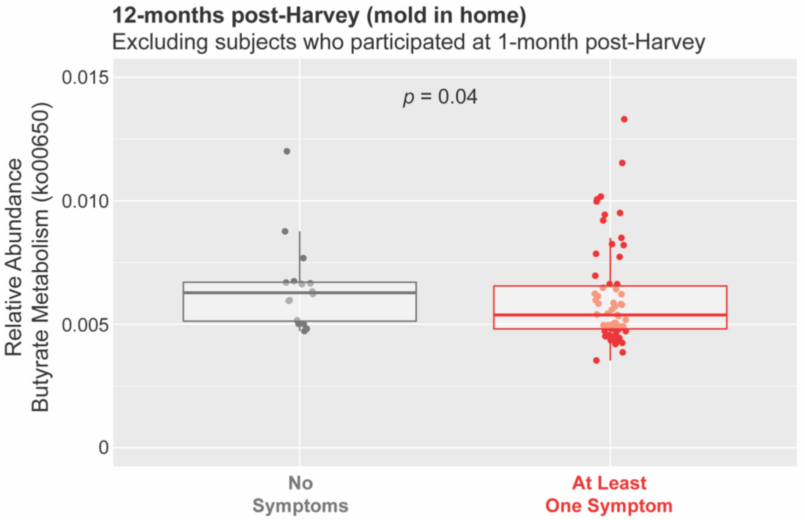


**Supplementary Figure 4. Relative abundance of butyrate metabolism genes excluding longitudinal participants.** Relative abundance of metabolic pathways involved in butyrate metabolism at 12-months post-Harvey (mold in home only), excluding subjects who participated at 1-month post-Harvey. (**c**, n=16 without symptoms, n=63 with symptoms, *p* =0.04, one-sided Mann-Whitney test). Box plot indicate median and interquartile range (IQR), whiskers show smallest (lower whisker) or largest (upper whisker) value within 1.5 times the IQR.

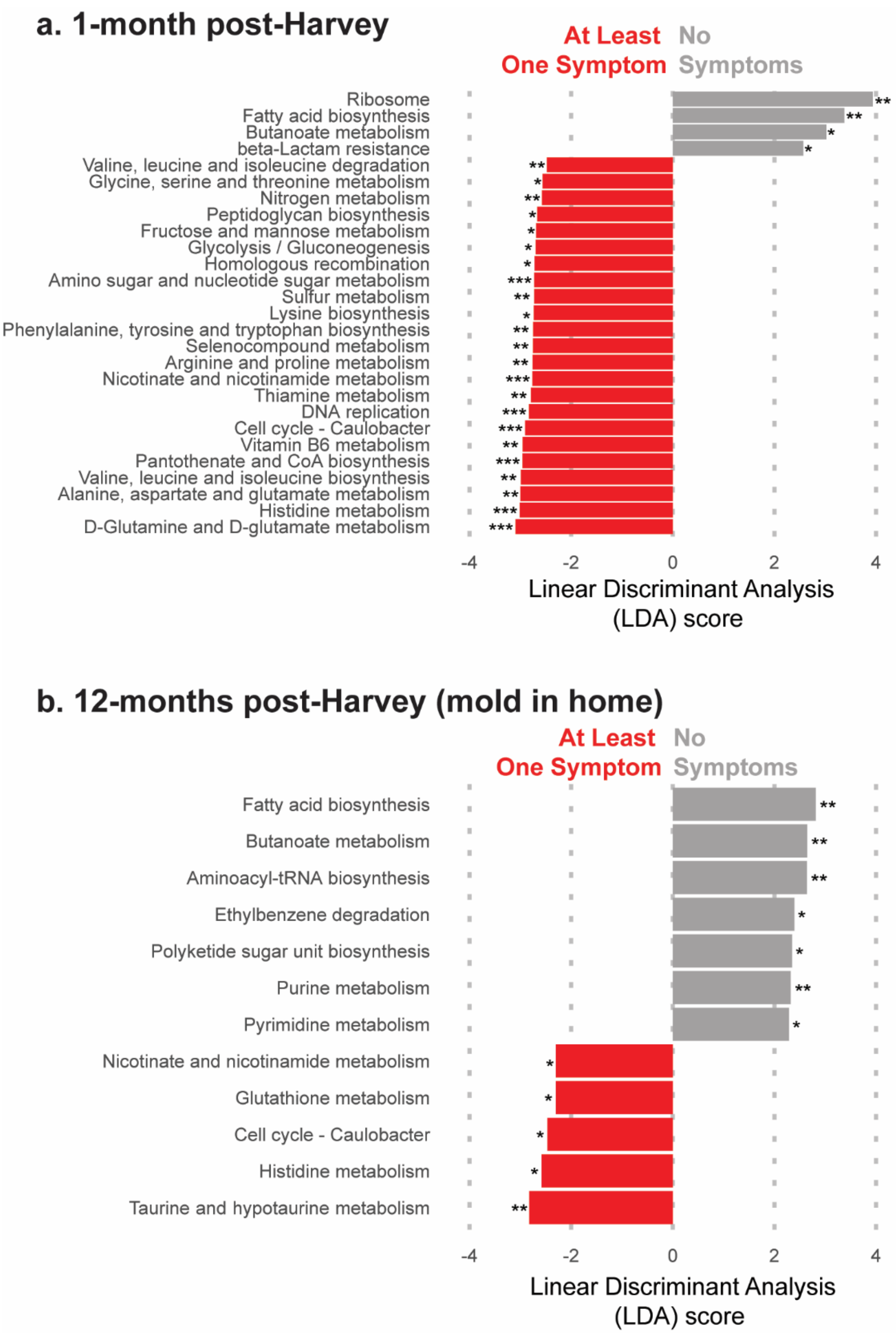


**Supplementary Figure 5. Gut bacterial metabolic pathways associated with allergic health outcomes.** Linear discriminant analysis Effect Size (LEfSe) reveals bacterial metabolic pathways profiled by WGS sequencing associated with absence of allergic symptoms (gray) and presence of allergic symptoms (red) at 1-month post-Harvey (**a**, n=18 without symptoms, n=88 with symptoms) and 12-months post-Harvey for subjects with signs of mold in the home (**b**, n=29 without symptoms, n=97 with symptoms) (LDA Score > 2.0, Kruskal-Wallis $p < 0.01$ (***), $p < 0.05$ (**), $p < 0.10$ (*)).

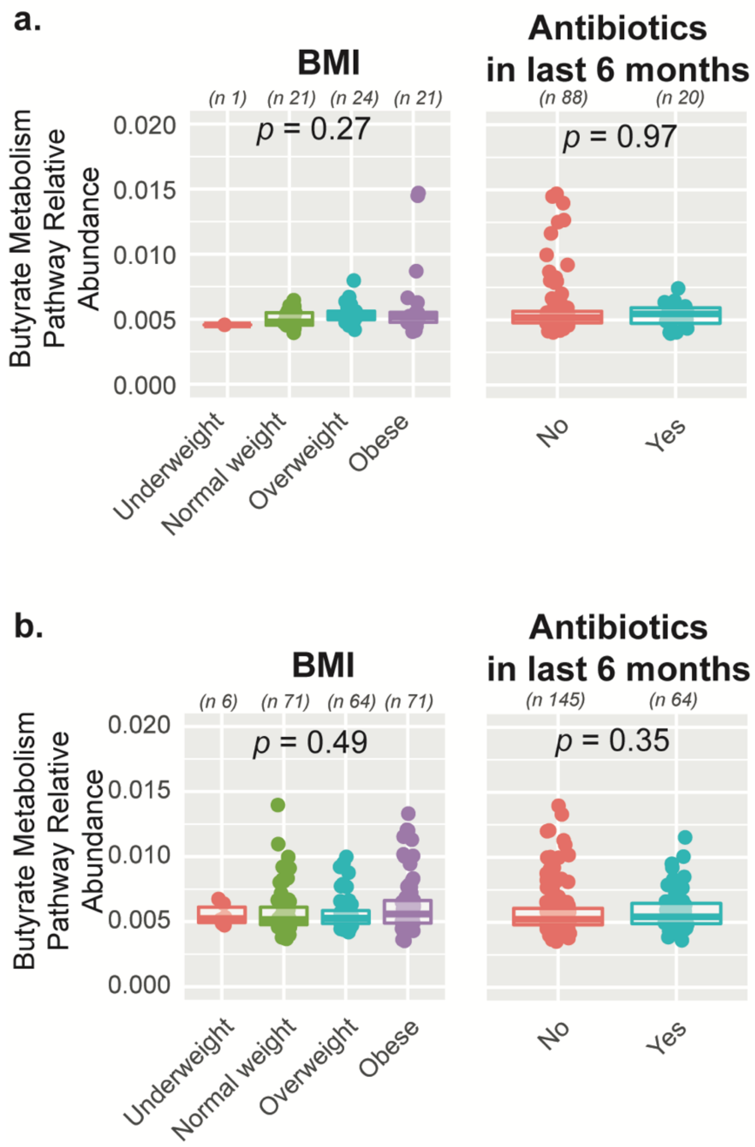


**Supplementary Figure 6. Variation in gut bacterial butyrate metabolism pathway (ko00650) abundance by BMI and antibiotic use.** Relative abundance of bacterial butyrate metabolism pathway in the gut microbiome at 1-month post-Harvey (**a**) and 12-months post-Harvey (**b**). Statistical tests between two groups are two-sided Mann-Whitney Tests, and statistical tests between more than two groups are Kruskall-Wallis tests. Box plots indicate median and interquartile range (IQR), whiskers show smallest (lower whisker) or largest (upper whisker) value within 1.5 times the IQR.

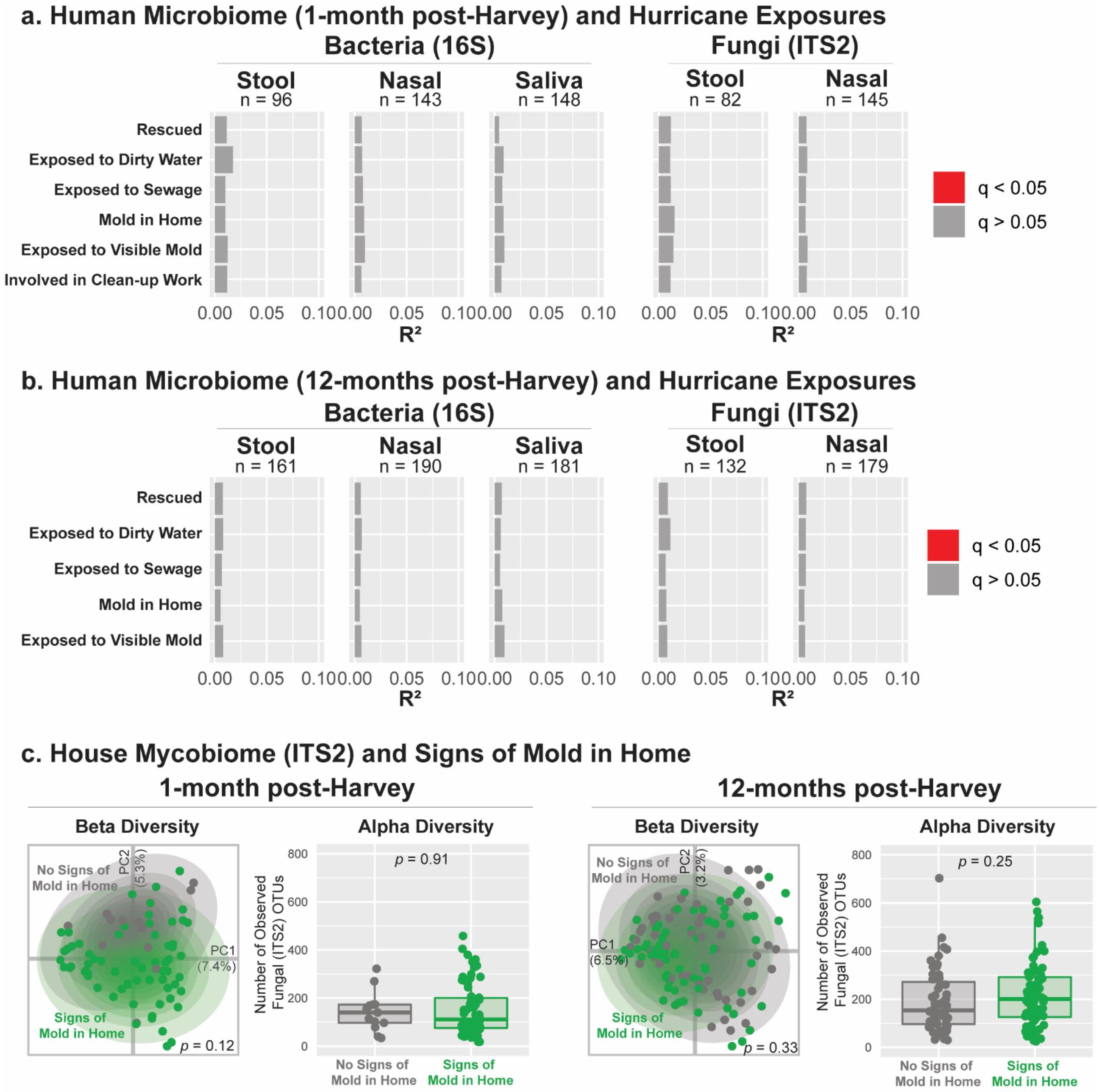


**Supplementary Figure 7. Relationship between hurricane exposures and fungal and bacterial microbiome composition.** PCoA of Binary Jaccard distances was used to evaluate associations between reported hurricane exposures and human microbiome composition at 1-month (**a**) and 12-months (**b**) post-Harvey. PERMANOVA was used to calculate $R^2$ (percent of total microbial variation accounted for by each variable) and p-values. Bonferroni correction of p-values was used to calculate q-values for each body site and time point. Sample numbers and statistics for each individual analysis are provided in Supplementary Table 4. **c.** Relationship between subject reported signs of mold in the home and house mycobiome (ITS2) beta and alpha diversity at 1-month post-Harvey (left, n=13 without signs of mold, n=64 with signs of mold) and 12-months post-Harvey (right, n=51 without signs of mold, n=73 with signs of mold). Beta diversity was calculated using PCoA of binary jaccard distances and significance of clustering was determined by PERMANOVA (1-month post-Harvey $p$ = 0.12 and $R^2$ = 0.015; 12-months post-Harvey $p$ = 0.33 and $R^2$ = 0.008); outer ellipse represents 95% normal confidence ellipse for each group. Alpha diversity was calculated as number of observed ITS2 Operational Taxonomic Units (OTUs) and two-sided Mann-Whitney tests were used to evaluate statistical significance. Only one sample per household was included in house mycobiome analyses. 12-month post-Harvey analysis only includes samples from subjects still living in the same home as they were at the time of Hurricane Harvey. Box plots indicate median and interquartile range (IQR), whiskers show smallest (lower whisker) or largest (upper whisker) value within 1.5 times the IQR.

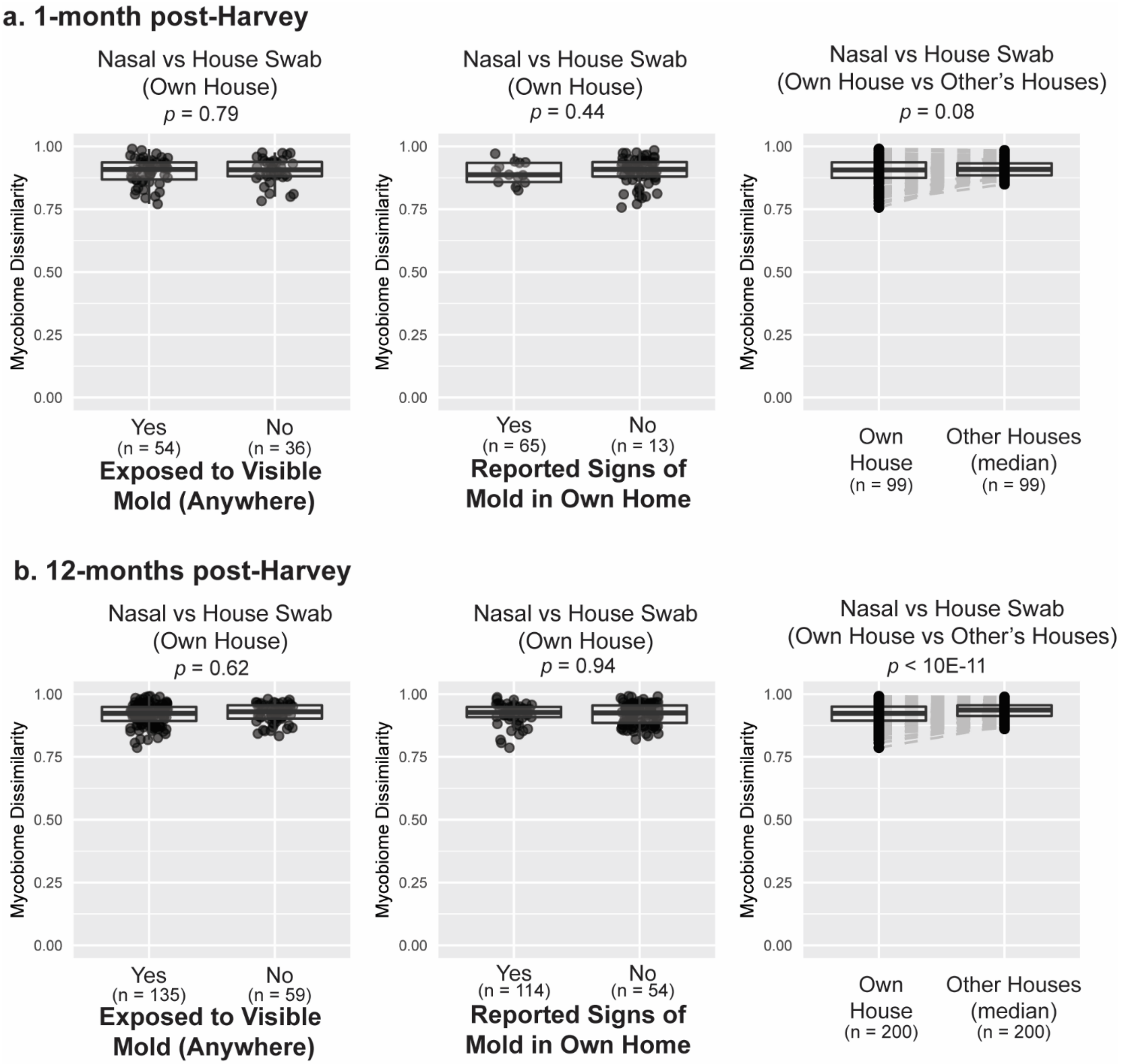


**Supplementary Figure 8. Mycobiome dissimilarity between nasal and household swabs.** Mycobiome (ITS2) dissimilarity (Binary Jaccard distance) analyses comparing the dissimilarity of each subject's nasal mycobiome to their own house mycobiome reveals that neither mold exposure (left) nor signs of mold in the home (middle) are associated with increased similarity between a subject's nasal mycobiome and their own household mycobiome 1-month (**a**) and 12-months (**b**) post-Harvey (two-sided Mann-Whitney test). Mycobiome dissimilarity analysis comparing each subject's nasal mycobiome to their own house mycobiome versus other subjects' house mycobiomes (median) reveals increased similarity between each subject's nasal mycobiome and their own home's mycobiome at 12-months but not 1-month post-Harvey (right, paired Mann-Whitney test). Box plots indicate median and interquartile range (IQR), whiskers show smallest (lower whisker) or largest (upper whisker) value within 1.5 times the IQR. N indicates the number of nasal vs. house dissimilarity distances included in each analysis.
